# Unsupervised co-optimization of a graph neural network and a knowledge graph embedding model to prioritize causal genes for Alzheimer’s Disease

**DOI:** 10.1101/2022.10.03.22280657

**Authors:** Vignesh Prabhakar, Kai Liu

**Affiliations:** gRED ECD AI, Genentech Inc., 1, DNA Way, South San Francisco, 94080, CA, USA

**Keywords:** Alzheimer’s Disease, Causal gene prioritization, Co-optimization, Protein-Protein interaction network, Knowledge Graph

## Abstract

Data obtained from clinical trials for a given disease often capture reliable empirical features of the highest quality which are limited to few studies/experiments. In contrast, knowledge data extracted from biomedical literature captures a wide range of clinical information relevant to a given disease that may not be as reliable as the experimental data. Therefore, we propose a novel method of training that co-optimizes two AI algorithms on experimental data and knowledge-based information from literature respectively to supplement the learning of one algorithm with that of the other and apply this method to prioritize/rank causal genes for Alzheimer’s Disease (AD). One algorithm generates unsupervised embeddings for gene nodes in a protein-protein interaction network associated with experimental data. The other algorithm generates embeddings for the nodes/entities in a knowledge graph constructed from biomedical literature. Both these algorithms are co-optimized to leverage information from each other’s domain. Therefore; a downstream inferencing task to rank causal genes for AD ensures the consideration of experimental and literature data available to implicate any given gene in the geneset. Rank-based evaluation metrics computed to validate the gene rankings prioritized by our algorithm showed that the top ranked positions were highly enriched with genes from a ground truth set that were experimentally verified to be causal for the progression of AD.

## 2. Introduction

Enriching empirical data collected from clinical studies/experiments with prior knowledge extracted from biomedical literature and updating the prior knowledge iteratively based on the new knowledge learnt from the experimental data is a novel approach of training in deep learning. The core objective behind this novel approach is to supplement the learning of an AI algorithm trained on experimental data with the learning of another AI algorithm trained on biomedical literature and vice versa so that both these algorithms could symbiotically benefit from each other’s data. We apply this novel training strategy to prioritize causal genes for Alzheimer’s Disease where one domain of data is a knowledge graph constructed from biomedical literature that relates clinical entities such as genes, diseases, signaling pathways, biological processes, molecular function, adverse events etc. [1][2] and the other domain of data is a protein-protein interaction (PPI) network [3–5] associated with experimental data that is derived from clinical studies such as common variant analysis [6–8], rare variant analysis [9–14], expression analysis [15–31] and gene knockouts from animal models [32–34] for Alzheimer’s Disease. In order to implement the same, we firstly enrich a graph neural network trained on the PPI network data with prior knowledge from a knowledge graph embedding (KGE) model trained on the biomedical knowledge graph. Thereafter, we update the prior knowledge in the KGE model using the node embeddings learnt by the graph neural network on the PPI network data for each epoch of training. This process is repeated iteratively to achieve co-optimization of the graph neural network and the KGE model. The node embeddings generated as a result of this co-optimization are used in a downstream inferencing task to rank all the causal genes relevant to Alzheimer’s Disease.

Alzheimer’s Disease is a neurodegenerative disorder that slowly destroys the memory and cognitive function of the patients diagnosed with it. The drug targets that have been successful in controlling the progression of the disease so far have been limited [35]. One of the key challenges in discovering novel drug targets for Alzheimer’s Disease has been the identification and prioritization of causal genes relevant to the disease [36]. Causal genes are the genes whose level of expression is directly associated with an increased risk for developing the disease. Causal genes are determined based on experimental genetic evidence available to implicate the genes through clinical studies such as common variant analysis, rare variant analysis, expression analysis, gene knockouts from animal models etc. as well as factual evidence from biomedical literature available to implicate the genes such as signaling pathway activation, molecular function, biological processes etc. Therefore, it becomes crucial to rank all the genes by causality in order to further experimentally explore the effect of gene regulation on the progression of Alzheimer’s Disease [37].

This can be achieved by assigning a unique score to each gene in the geneset based on the experimental data and knowledge-based information from literature that is available to determine if a given gene is causal to the risk of developing Alzheimer’s Disease. This is known as gene scoring [38]. Gene scoring is a crucial step in the process of target qualification in order to identify and rank the causal genes relevant to Alzheimer’s Disease. Genes can be scored using a scoring function that accepts the experimental data and factual information from literature for each gene as the inputs to yield a unique score for each gene which can then be sorted to rank the genes based on their causality to Alzheimer’s Disease. The factual information relevant to each gene such as signaling pathway activation, biological processes and molecular functions is derived from a biomedical knowledge graph (Hetionet) [1][2]. The experimental data contains features derived from common variant analysis, rare variant analysis, expression analysis and gene knockouts from animal models for each gene in the geneset. Furthermore, the multiple genes participating in coding a single protein are closely associated and linked with each other in the protein-protein interaction network. Therefore; the experimental data relevant to each gene can be diffused as node features in the protein-protein interaction network to help the graph neural network algorithm in associating the node features of all the connected genes. The scoring function used for assigning a unique score to every gene in the geneset is the objective function learnt by our co-optimization algorithm that leverages information from the knowledge graph to supplement the information learnt from the experimental data and vice versa. The ground truth that we used for testing and validating the gene rankings prioritized by our algorithm is a set of silver-standard genes that were collected from expert curated data sources such as Drugbank, ChEMBL, Opentargets, OMIM etc. and have verified causal evidence implicating them with Alzheimer’s Disease [39–42].

Rank based evaluation metrics [43] showed significant improvement for the silver standard genes in the geneset prioritized by co-optimizing the knowledge graph embedding model with the graph neural network model.

## 3. Literature Review

There have been previous approaches that have utilized graphs/networks along with experimental genetics data to prioritize causal genes so that they could be explored as potential drug targets for a given disease. The first such approach was adopted by the GWAB algorithm [44] that prioritized candidate genes by network-based boosting of GWAS (Genome wide association studies) data. It leveraged networks to analyze GWAS summary statistics [45] to prioritize top genetic variants associated with a disease and aimed to detect weakly implicated genes via disease-gene associations in a human gene functional interaction network called HumanNet [46] by their proximity to other strongly implicated genes in a molecular network using a Naive Bayes guilt-by-association algorithm [47]. Similar to the GWAB algorithm; we also use two types of graphs/networks in our proposed algorithm so as to associate the genetic evidence relevant to causal genes from a protein-protein interaction network that takes experimental data as node features with the functional evidence such as disease-gene associations from a biomedical knowledge graph. Although the GWAB algorithm picked weakly implicated genes from a human gene functional interaction network and used that information to measure their proximity to strongly implicated genes in a molecular network; it did not use any representation learning algorithm (such as a graph neural network or knowledge graph embedding model) to derive those insights and therefore could have easily missed out on selecting weakly implicated genes that did not have a direct disease-gene connection in the functional interaction network and also could have easily missed out on measuring the proximity of the weakly implicated genes with several other strongly implicated genes that did not have a direct connection with the weakly implicated genes in the molecular network. This is precisely the problem that a graph neural network solves by generating embeddings/vectors to represent the underlying latent semantic structure of a graph/network that can be missed by a simpler algorithm that only considers the most apparent direct connections in the graph during inferencing [48]. Furthermore, GWAS summary statistics only implicates common genetic variants for a disease and misses out on considering other significant modalites of genetic evidence such as implication of rare genetic variants, differential gene expression and baseline expression as well as gene knockouts from animal models.

Another algorithm that has been proposed in literature to prioritize the top genetic variants associated with a disease using GWAS summary statistics and network data is the NetWAS algorithm [49]. This algorithm combined genes with nominally significant GWAS P-values and tissue-specific networks [50] to identify disease-gene associations more accurately than GWAS alone. Each tissue network represented the tissue-specific posterior probability of a functional relationship between each pair of genes. NetWAS applied support vector machines [51] using nominally significant P-values as positive examples and randomly selected genes as negative examples to construct a classifier that identified tissue-specific network connectivity patterns associated with the hypertension phenotype. The features of the classifier were the edge weights of the labeled examples of all the genes in the network. Genes annotated to hypertension phenotype in the Online Mendelian Inheritance in Man (OMIM) database [42] were more highly ranked by this classifier than by the initial GWAS and also found antihypertensive drug targets from Drugbank [39], Therapeutic Target Database (TTD) [52] and Comparative Toxicogenomics Database (CTD) [53] that were more enriched by NetWAS than by GWAS. Although this approach was able to combine common variant statistics (nominally significant GWAS P-values) for candidate genes with tissue-specific networks in order to identity disease-gene associations; it did not consider any other significant modalites of genetic evidence such as implication of rare genetic variants, differential gene expression and baseline expression as well as gene knockouts from animal models etc. Additionally, the connectivity in tissue-specific networks is often deficient and therefore fails to capture functional information relevant to other useful associations for a given phenotype.

Prioritizing target-disease associations with novel safety and efficacy scoring methods evaluates the efficacy and safety of potential drug targets by proposing efficacy scores that utilize existing gene expression data and tissue/disease specific networks to improve the inferencing on target-disease associations [54]. Although this approach considered differential gene expression for implicating causal genes it did not consider other modalities of crucial genetic evidence such as implication of causal genes by common genetic variants, rare genetic variants etc. Differential expression data is not sufficient by itself to evaluate the efficacy and safety of potential drug targets. Furthermore, the safety of a druggable target depends on the adverse events associated with it. The approach that we have proposed in this paper obtains the adverse event data from the knowledge graph that relates several pieces of factual information such as genes, diseases, signaling pathways, biological processes, molecular functions, adverse events etc. It is therefore more informative and relevant in assessing the efficacy and safety of potential drug targets over a tissue/disease specific network.

The PoPs algorithm leverages polygenic enrichments of gene features to predict genes underlying complex traits and diseases using a similarity based gene prioritization method [55]. The PoPs algorithm uses MAGMA eval (linear regression) of GWAS data [56] to train another linear regression model that uses functional annotations such as baseline gene expression from gTEX [15], biological pathways and predicted protein-protein interaction data as the gene features. PoPs avoided benchmarking using curated silver standard genesets that may be biased towards well-studied genes or genes in well-characterized pathways. Instead, PoPs used a slightly different approach for benchmarking by estimating the average contribution of SNPs in genes with high priority scores per SNP heritability [57]. Thereafter, assuming that the causal gene is often the closest gene to the lead variant in the locus, PoPs tested whether the prioritized genes were more often the closest gene to the lead variant in the locus than expected by chance. The PoPs algorithm was a promising approach to prioritize causal genes but the underlying linear regression algorithm utilized very few functional annotations / gene features to rank causal genes. Experimental data such as the evidence from common variant and rare variant studies were not considered when generating the gene features for the linear regression model. Moreover, we do not know if the relationship between the gene features and gene scores can be modeled using a linear function and the algorithm may underfit the training data leading to poorly prioritized causal genes.

There have been previous studies that have integrated functional genomics and immune-related annotations, together with knowledge of network connectivity in order to maximize the informativeness of genetics for target validation [58]. This approach has adopted a priority index (PI) pipeline that takes genome-wide association study (GWAS) variants for specific immune traits as inputs and defines seed genes using genomic predictors such as distance to SNP [59], physical interaction, gene expression regulation, EQTL colocalization [60] etc. to identify and score the genes that are likely responsible for the GWAS signals. Additional scores for annotation predictors such as immune function (fGene), immune phenotype (pGene) and rare genetic diseases related to immunity (dGene) are then only applied to the seed genes. Thereafter, random walk is performed on the STRING protein-protein interaction (PPI) network [3] in order to identify non-seed genes that lack genetic evidence but are highly ranked based on network connectivity alone and also to enhance the scoring for the seed genes with evidence of network connectivity. This study notably found that the interacting neighbors in a PPI network tend to be known drug targets rather than the GWAS reported genes. The priority index pipeline was evaluated by extracting existent drug therapeutics and target genes from the ChEMBL database [40] and comparing those against the ones prioritized by the priority index pipeline.

Disease module identification based on representation learning of complex networks integrated with GWAS, eQTL Summaries and Human Interactome derived disease related modules from an integrated network with multi-layer information including human interactome (mainly protein-protein interactions) and summaries of GWAS and eQTL studies [61]. This approach leverages a community detection algorithm called N2V-HC to learn node representations in a molecular network and unbiasedly detect gene communities enriched with potential disease genes. The idea behind this community detection algorithm is driven by the primary observation that disease-related proteins tend to interact closely in biological networks and that disease-related proteins tend to form many separate connected components which are scattered across the network [62]. Thereafter, a Fischer test [63] is used to test whether the causal genes for a disease are enriched in the candidate disease module. The N2V-HC algorithm is a promising approach that groups/clusters a network based on causal gene communities for a given disease. We adopt a similar approach in our algorithm for grouping the genes into separate clusters using the cosine similarity between the node representation of a given gene *G*_1_ and the node representation of a causal, silver standard gene and assign the cosine similarity score as the gene score for *G*_1_. Other than the evidence incorporated from common and rare variant studies it will also be useful to incorporate functional/factual data relevant to each gene in the geneset. The factual data could include relevant pathways, molecular functions, biological processes, cellular components etc. for a given gene *G*_1_

Rosalind is a gene prioritization method that combines heterogeneous knowledge graph construction with relational inference via tensor factorization to accurately predict disease-gene links in the graph [64]. It uses a subgraph consisting of Disease-GeneProtein links with the ‘Therapeutic Relationship’ relation type as a benchmark. The tensor factorization model is trained on the full knowledge graph and evaluated on the subgraph consisting of the Disease-GeneProtein links and the ‘Therapeutic-relationship’ relation type [65]. Disease-Disease and Compound-Compound relationships are not included, as the former is not available across enough of diseases of interest while the latter is not available for measures of functional similarity at sufficient resolution. Knowledge graphs are generally enriched with functional/factual data and can therefore prioritize causal genes solely based on the factual information. However, experimental data from common variant analysis studies (GWAS), rare variant analysis studies (burden tests) and differential gene expression analysis studies are extremely crucial to prioritize casual genes and cannot be excluded when designing an algorithm for the same purpose. Both factual data and experimental data may be independently insufficient in prioritizing causal genes but aggregating them together gives the algorithm/model multiple feature attributes to score all the genes in the geneset.

Finding the targets of a drug by integration of gene expression data with a protein-protein interaction network proposes a network-based computational method for drug target prediction, applicable on a genome-wide scale [66]. This approach relies on the analysis of gene expression following drug treatment in the context of a functional protein association network. By diffusing differential expression signals to neighboring or correlated nodes in the network, genes are prioritized as potential targets based on the transcriptional response of functionally related genes. AUC values of up to 90% demonstrated the effectiveness of this approach and indicated the predictive power of integrating experimental gene expression data with prior knowledge from protein-protein interaction networks. The main idea behind our algorithm takes inspiration from this study as we also attempt to incorporate prior knowledge from a knowledge graph in the training procedure of a graph neural network on a protein-protein interaction network diffused with experimental data from common variant studies, rare variant studies, expression analysis studies and gene knockouts from animal models. The difference between these approaches is primarily in terms of the different modalities of data that have been used for training the models and the novel training procedure (co-optimization) that we have adopted to incorporate the prior knowledge. While this study integrates prior knowledge from a protein-protein interaction network with experimental gene expression data; our approach integrates prior knowledge from a knowledge graph into a protein-protein interaction network that is diffused with experimental data. This is a promising approach to rank causal genes which can be further improved by diffusing more experimental data features other than gene expression into the protein-protein interaction network. A combination of common variant analysis features, rare variant analysis features and gene expression analysis features is required in terms of the experimental data needed to prioritize causal genes.

Drug target prioritization by perturbed gene expression and network information also proposed a network-based computational method for drug target prediction, applicable on a genome-wide scale but showed that the gene expression of drug targets is usually not significantly affected by the drug perturbation and therefore expression changes after drug treatment on their own are not sufficient to identify drug targets [67]. However, ranking of candidate drug targets by network topological measures prioritizes the targets. In order to demonstrate the same, they introduce a novel method called local radiality that combines perturbed genes and functional interaction network information and this new method outperforms other methods in target prioritization including a random walk approach [68] and proposes cancer-specific pathways from drugs to affected genes. Like the other network-based computation methods explored for drug target prediction this method also leverages limited experimental features and abundant functional interaction features from the networks. Therefore; this method also needs to incorporate features from common variant studies and rare variant studies to achieve reliable results for ranking causal genes that can be explained experimentally.

## 4. Dataset

The primary objective of our methodology is to incorporate learning from multiple domains of data and augment the learning from one domain with the learning from the other domain. Keeping this in mind we carefully selected the datasets accordingly. The datasets that we have leveraged for training our algorithm fall broadly into the following categories:

1. Experimental genetics data
2. Protein-Protein interaction network
3. Biomedical knowledge graph (Hetionet)
4. Silver standard genesets relevant to Alzheimer’s Disease (Ground truth set)
5. Geneset to be prioritized

### 4.1 Experimental genetics data

We consolidated experimental genetics data for incorporating causal gene evidence relevant to Alzheimer’s Disease from four different experimental sources namely common variant analysis, rare variant analysis, expression analysis and gene knockouts from animal models. The features in the experimental genetics data are available for each individual gene present in the geneset that we try to rank using our algorithm.

#### 4.1.1 Common variant analysis

A common variant by definition is a variant whose allele frequency is greater than one percent. Common variants are easy to measure and help us study complex traits via genome wide association studies (GWAS) [6–8]. GWAS typically reports only variant-level data and does not provide gene-level mapping for the variant-level data. Therefore, we built a system to link variants to genes. Common variants are common in a population and are likely to either have been subjected to selective pressure or undergone neutral evolution over an extended period of time. GWAS helps us find regions of the genome associated with complex traits by testing individual variants across the genome.

We study common variants for the following reasons:

- Common variants lie in intergenic regions and are thought to play regulatory roles in gene function.
- Common variants are easy to measure and help us study complex traits.

The data obtained from common variant analysis includes the following set of features for each individual gene in the geneset:

- Aggregated gene p-values from GWAS study variants (calculated by PASCAL).
- Posterior probability for colocalization of eQTL and GWAS signals for a gene (calculated using coloc).
- Presence/absence of a missense mutation in the gene’s coding region of proteins.

#### 4.1.2 Rare variant analysis

Rare variants are those whose allele frequency is less than one percent, are exonic, and functionally relevant. These tend to be either novel mutations in a population that have not yet been subjected to significant selective pressure or older variants kept at lower frequencies due to their deleteriousness. [9–14]

We study rare variants for the following reasons:

- They can help identify genes that impact a disease or phenotype
- For some common diseases, common variants do not sufficiently explain the genetic basis of disease.

The data obtained from rare variant analysis includes the following set of features:

- Rare variant p-values obtained from burden test.
- Burden test odds ratio

#### 4.1.3 Expression analysis

We collect expression data for each gene using expression analysis. Expression analysis falls into two categories:

- Baseline Expression from Genotype-Tissue Expression (GTEx) [15]
- Differential Expression Analysis [16–31]

- Bulk RNA Sequencing and
- Single-Cell RNA Sequencing (scRNA)

#### 4.1.4 Gene knockouts from animal models

We also incorporate gene knockout scores obtained from clinical trials in our experimental genetics data. Knockouts are collected by downregulating the expression of a particular gene or fully eliminating it in animal models such as mouse, zebrafish and fruit-fly [32–34]. Observing the progression of Alzheimer’s Disease by regulating the level of expression of the genes in animal models helps us identify if those genes are potentially causal in nature.

### 4.2 Protein-Protein interaction network

We consolidate three different networks to build a unified protein-protein interaction (PPI) network namely StringDB, Bioplex and Immunoglobulin Superfamily (IgSf) [3–5]. The consolidated protein-protein interaction network is a homogeneous graph that has 20,118 nodes and 1,211,771 edges. Each node in this network is a unique gene and edges between two gene nodes exist if they are involved in coding the same protein. The features in the experimental data are passed as node features / attributes to the consolidated protein-protein interaction network. This is done so that our algorithm takes into consideration both the experimental data as well as graph connectivity from the protein-protein interaction network while generating vector representations / embeddings for our input data.

### 4.3 Biomedical knowledge graph

Biomedical knowledge graph is a heterogeneous knowledge graph that relates several diverse clinical entities such as genes, diseases, signaling pathways, biological processes, molecular functions, adverse effects etc. [69]. They are constructed by extracting information from literature and other expert curated datasets. One such biomedical knowledge graph is Hetionet [1][2]. Hetionet contains 47,031 nodes of 11 types and 2,250,197 edges of 24 types. We use hetionet to generate the gene scores based on the evidence available in literature relevant to Alzheimer’s Disease. Embeddings/vector representations can be generated for the nodes and edges in the knowledge graph using a knowledge graph embedding (KGE) model. These embeddings can in turn be used downstream to rank the relevant genes based on the likelihood of existence of a link between the gene nodes and the Alzheimer’s Disease node in the knowledge graph (Hetionet).

### 4.4 Silver standard genesets relevant to Alzheimer’s Disease

Due to the absence of any ground truth sets; we curated a set of 400 genes relevant to Alzheimer’s Disease that have been previously explored as potential drug targets and they serve as the semi ground truths for our algorithm. We refer to it as the silver standard geneset. These were primarily created from four different expert-curated, open-sourced data repositories namely:

1. OpenTargets [41]
2. DrugBank [39]
3. ChEMBL [40]
4. Online Mendelian Inheritance in Man (OMIM) Database [42]

### 4.5 Geneset to be prioritized

We collected a set of 20,000 genes to form a geneset. Each gene in this geneset possesses experimental data collected from clinical studies and knowledge-based factual data collected from biomedical literature. The objective of our algorithm is to prioritize/rank all the 20,000 genes in this geneset based on the evidence available to determine if their regulated expression is causal in nature to developing Alzheimer’s Disease.

## 5. Methodology

Our objective is to define a scoring function that assigns a score for each individual gene based on which we can rank all the genes in the geneset. One component of the scoring function consumes the features from experimental genetics data for each gene along with its relevant connectivity information from the consolidated PPI network to yield a score *S*_1_. This score *S*_1_; could be further utilized to rank all the genes in the geneset. The other component of the scoring function consumes the nodes and edges from the knowledge graph as input to yield a score *S*_2_ for each gene in the geneset. Thereafter, we aggregate both these components of the scoring function by averaging *S*_1_ and *S*_2_ to generate a single set of scores, *S_agg_*, for each gene in the geneset as shown in equations 1, 2 and 3.

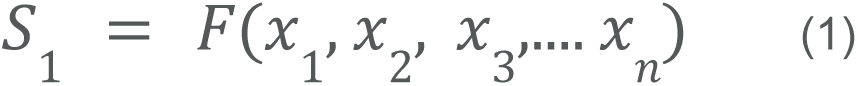

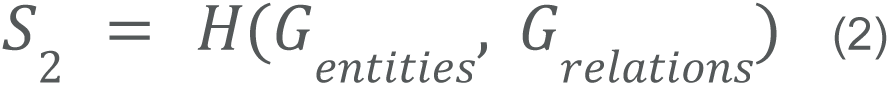

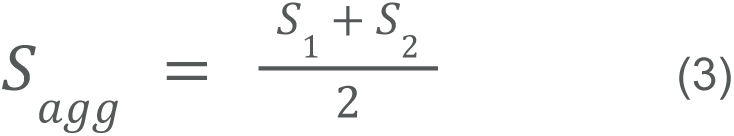

As shown in Figure 1, we adopt a divide and conquer approach [70] to design both the components of the scoring function. We leverage two different modeling techniques for two different modalities of data. We use a graph neural network algorithm called Deep Graph Infomax [71] to generate scores for all the genes in the geneset using the experimental genetics dataset and the protein-protein interaction network and we use a knowledge graph embedding model to generate scores for the same set of genes in the geneset using the knowledge graph (Hetionet). Furthermore; we co-optimize both these algorithms so that the learning outcome from the knowledge graph embedding model (Implication of causal genes based on evidence in literature) supplements the learning outcome from the deep graph infomax algorithm (Implication of causal genes based on experimental evidence) and vice versa. Therefore, the resultant scoring components S_1_ and S_2_ generated by the deep graph infomax model and the KGE model respectively are mutually informative in nature as a result of this co-optimization.

**Figure 1:**
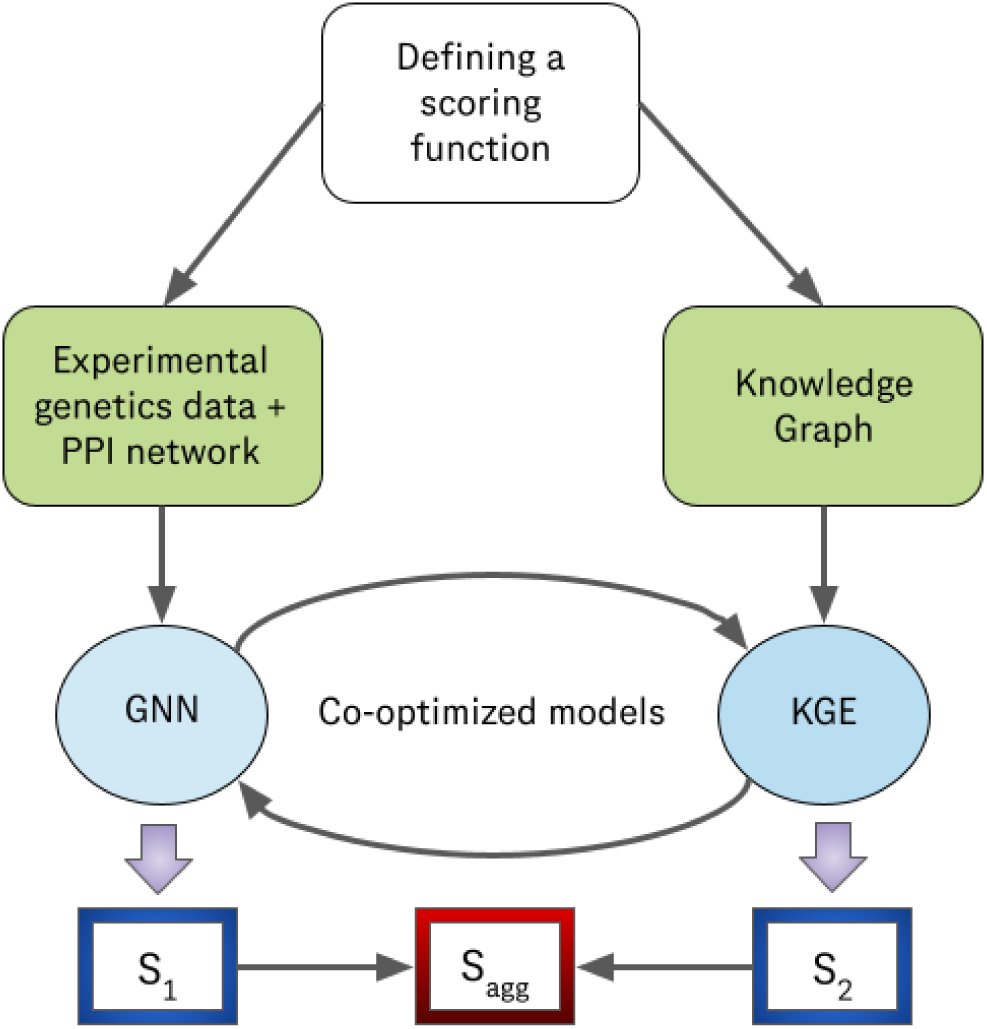
Divide and conquer strategy to design a scoring function that ranks the genes relevant to Alzheimer’s Disease using two different domains of data - experimental genetics data incorporated into a PPI network and factual/literature data from a biomedical knowledge graph.

### 5.1 Graph Neural Network: The Deep Graph Infomax algorithm

The deep graph infomax algorithm is a graph neural network algorithm used to generate unsupervised embeddings for the nodes in a graph [71]. It uses a graph neural network encoder to generate node embeddings for the actual graph and a corrupted graph generated from the actual graph. Thereafter it distinguishes between the node embeddings derived from the true graph and the node embeddings derived from the corrupted graph and in the process understands the sensible connections possible for a given node.

#### 5.1.1 Generating unsupervised node embeddings for the protein-protein interaction network augmented with the experimental genetics data passed as node features to the network

Figure 2 shows the architecture of the deep graph infomax algorithm that we leverage to produce unsupervised node embeddings for the protein-protein interaction (PPI) network shown in Figure 3 that is diffused with experimental genetics data as its node features.

**Figure 2:**
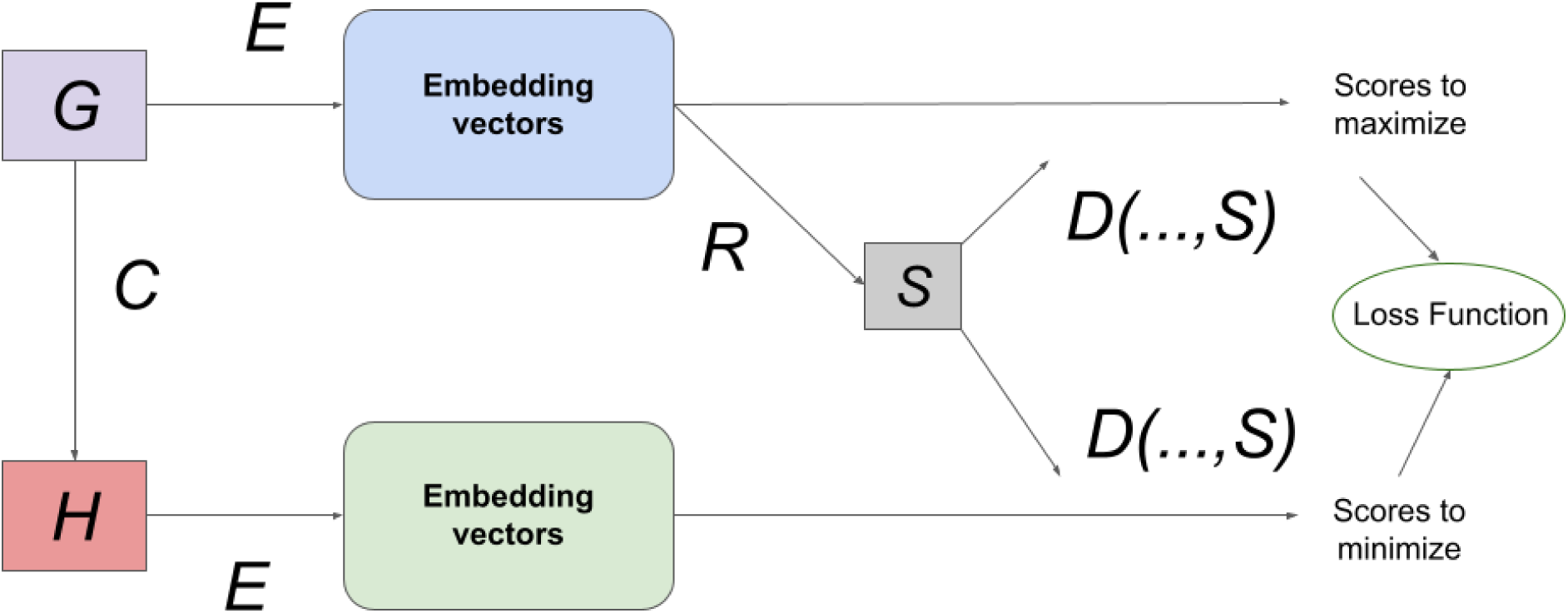
The architecture diagram of a deep graph infomax algorithm used to generate unsupervised embeddings/vector representations for the gene nodes and their respective experimental genetic features in the protein-protein interaction network.

**Figure 3:**
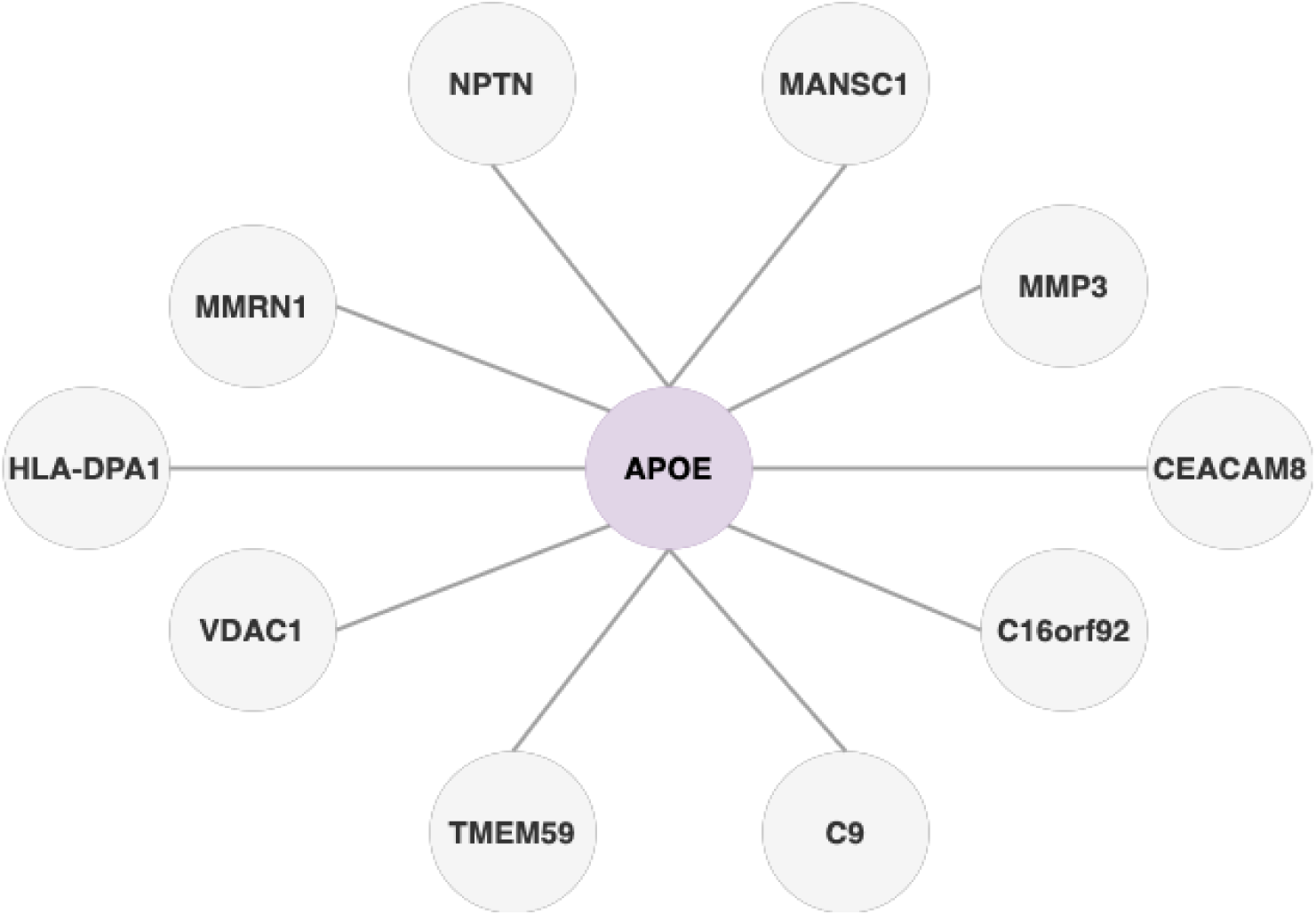
A protein-protein interaction network is a graph where each node is a gene and an edge exists between two or more genes if they are a part of the coding region of the same protein. The experimental genetics data associated with each gene is passed as the gene’s node feature in this protein-protein interaction network.

The Deep Graph Infomax algorithm generates node embeddings in an unsupervised manner. We take the PPI network *G* and corrupt it by randomly shuffling the node features among the gene nodes in the graph to create a mutated graph *H*. *H* contains the same connections as *G* but the node features associated with each individual gene node differ in comparison to the node features associated with each individual gene node in the true PPI network *G*.

We then use an encoder to create vector representations / embeddings for the nodes in both the true PPI network *G* as well as the mutated PPI network *H*. The encoder *E* is a graph neural network algorithm that represents latent information from the semantic structure / topology of the graph in the form of embeddings. We use a GNN Encoder (graph convolutional network (GCN) / graph attention network (GAT) / GraphSAGE / APPNP) [72–75] to encode the connectivity between the nodes along with their respective node features in both the true PPI network *G* as well as the mutated PPI network *H*. We use a readout mechanism *R* to summarize the node embeddings from the true PPI network. This is done by simply averaging the node embeddings from *G* to create a graph embedding vector *M* for *G*. A score *S* is assigned to each node embedding vector using a discriminator *D* that accepts a node embedding vector *H* from either *G* or *H* as input along with the summarized graph embedding vector *M*. Thereafter, the scores are collected and combined in a loss function that tries to maximize *S* if *Y_i_* is a node embedding vector from *G* and minimize *S* if *Y_i_* is a node embedding vector from *H*. Post optimization; the discriminator yields scores closer to 1 for the node embeddings from the true PPI network *G* and scores closer to 0 for the node embeddings from the mutated PPI network *H*. The loss function to be optimized for generating unsupervised node embeddings is shown in equation 4.

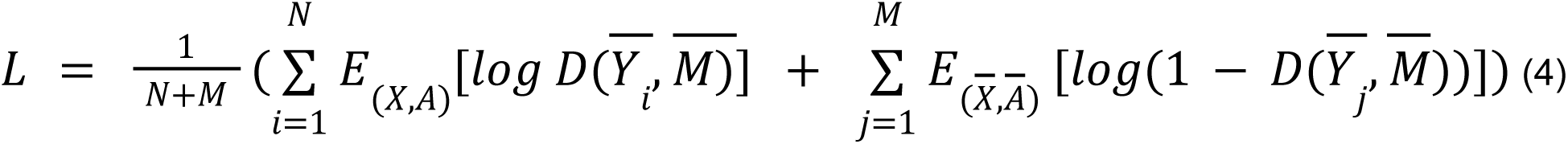

#### 5.1.2 Downstreaming strategy to generate gene scores

Once the deep graph infomax algorithm generates node embeddings for the gene nodes in the PPI network we obtain the node embeddings for both silver standard genes as well as non silver standard genes. Next, we want to group the non silver standard genes with silver standard genes. This is done to identify the closeness/proximity of the non-silver standard genes to the silver standard genes in terms of experimental evidence. To achieve this we take cosine similarity of the embeddings of a non silver standard gene with that of the embeddings of every silver standard gene. Then we assign the maximum cosine similarity score obtained with a particular silver standard gene as the gene score of the non silver standard gene [76]. Likewise, we assign a gene score (cosine similarity score) to all the genes in the geneset that lie in the interval (0,1). This number indicates the closeness between a non silver standard gene and a silver standard gene that has been historically explored as a potential drug target for Alzheimer’s Disease in terms of the experimental features in our dataset.

### 5.2 Knowledge graph embedding model

A knowledge graph embedding (KGE) generates embeddings / vector representations that capture latent properties of the nodes and edges in a graph [77]. These embeddings can then be used in downstream machine learning tasks such as link prediction [78]. In general, the likelihood of existence of a link between two nodes in the KG can be predicted by computing the proximity of a head node embedding and edge embedding with that of a tail node embedding by passing them as inputs to model-specific scoring functions as shown in Table 1 and computing a plausibility score for the existence of a link between the two nodes [79]. This can be repurposed to rank a set of candidate tail nodes for a given head node and edge. Therefore, we can rank a set of candidate gene nodes and generate gene scores for those gene nodes based on their proximity to the Alzheimer’s Disease node in the knowledge graph using the same link prediction strategy outlined above.

**Table 1.**
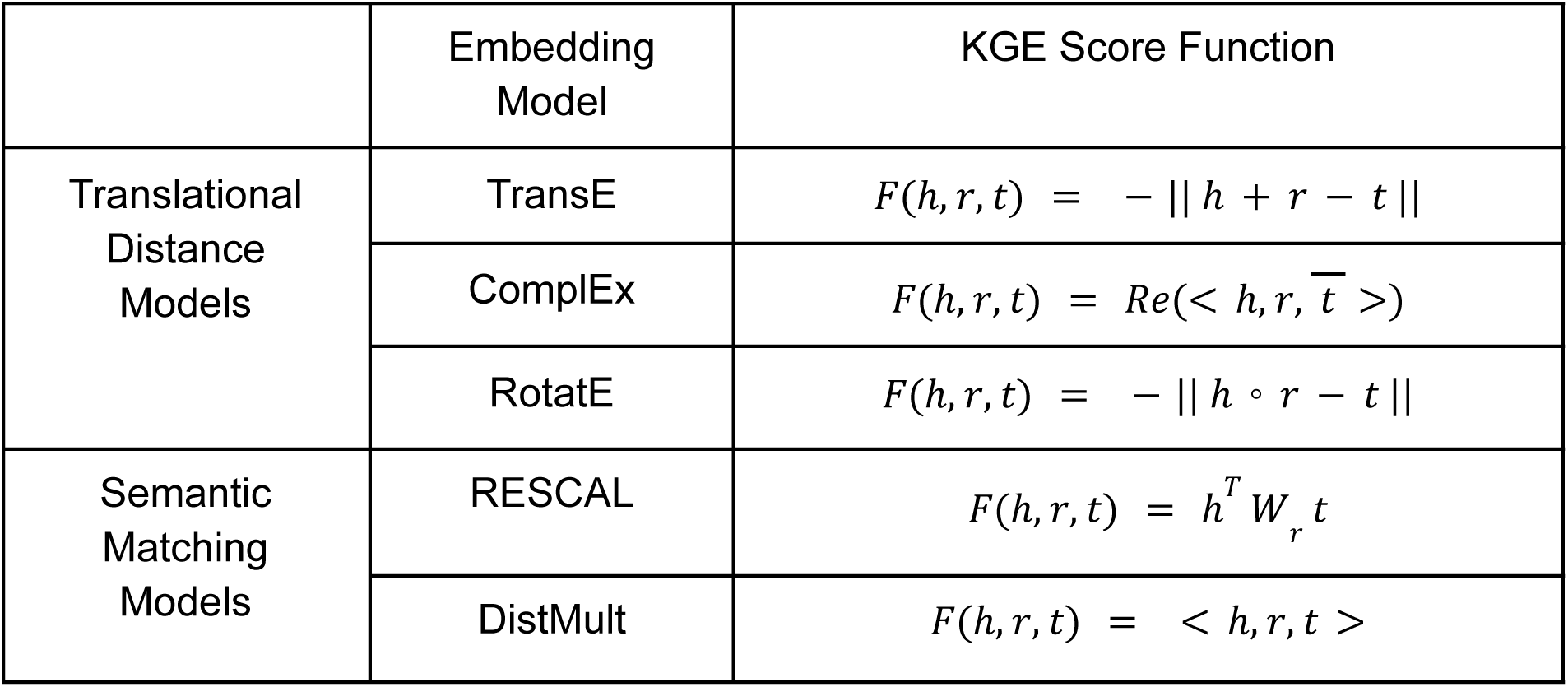
Scoring functions of standard knowledge graph embedding models given the head entity vector representation h, tail entity vector representation t and relation vector representation

#### 5.2.1 Generating vector representations/ embeddings for the entities and relations in Hetionet

A knowledge graph (KG) is a collection of known facts represented in the form of a directed labeled heterogeneous graph, wherein each node represents an entity and each edge represents a relation between the entities. Each fact is represented in the form of a triple (head node, relation, tail node). For example, the fact that Amyloid β is a signaling pathway of Alzheimer’s Disease can be stored in the form of a triple as (‘Amyloid β’, ‘pathway_of’, ‘Alzheimer’s Disease’) where Amyloid β and Alzheimer’s Disease are the nodes / entities and *pathway_of* is the edge / relation that connects them together. The categories or classes of entities and relations in the knowledge graph are standardized to a closed set. Hetionet is a knowledge graph constructed from biomedical literature that relates a diverse set of clinical entities that includes genes, diseases, signaling pathways, molecular functions, biological processes, cellular components, symptoms, adverse events / side effects, compounds / drugs etc. [1][2] as shown in Figure 4. It is open-sourced and has about 2.2M triples. These clinical entities are extracted from literature as well as other expert-curated sources of structured data and are useful in ranking genes to explore them as potential novel drug targets for a particular disease.

**Figure 4:**
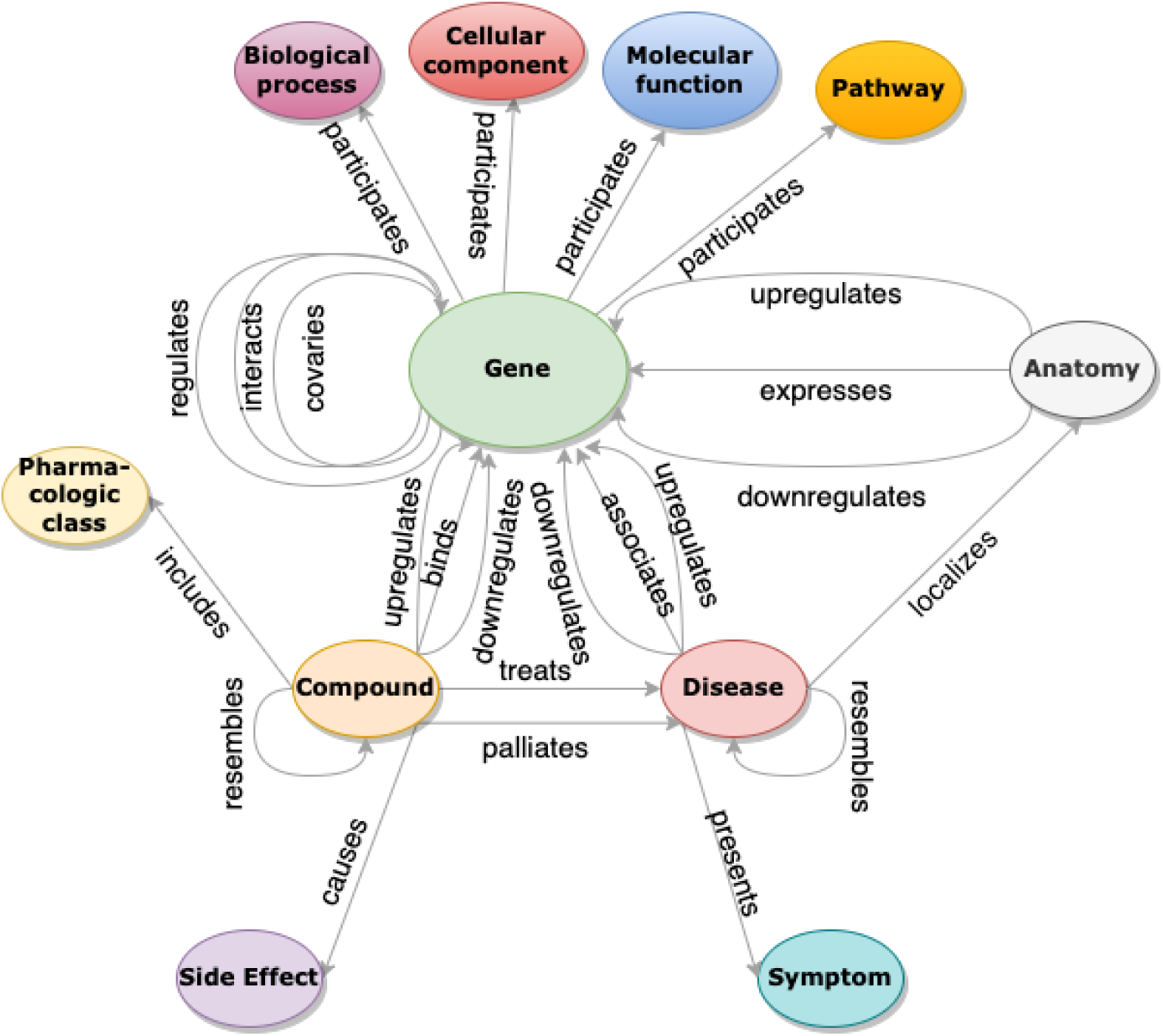
Hetionet is a knowledge graph that extracts heterogeneous facts from literature relevant to all the genes in the geneset such as signaling pathway, biological process, molecular function, cellular component, pharmacologic class etc. and links them together using nodes and edges. The knowledge graph thereby contains important prior knowledge that is absent in the experimental genetics data.

The other component of our scoring function to rank the genes comes from the knowledge graph embedding model (KGE). A knowledge graph embedding (KGE) model is an algorithm that generates embeddings / vector representations of the nodes and edges to capture latent semantic properties of the entities and relations in the KG from its structure / topology. Knowledge graph embedding models generate a given number of negative triples (synthetic triples) for every positive triple (true triples) that exists in the KG using a negative sampler [80]. This is done to help the KGE model distinguish between the nodes that can be connected for yielding a plausible triple that can exist in the KG and the nodes that upon connection would yield a triple that is unlikely to exist in the knowledge graph using a suitable loss function. Knowledge graph embedding models are optimized using a margin ranking loss function [81][82]. It is a linear-to-rank loss that is used for maximum-margin classification in pairwise settings to distinguish between positive triples ^+^ and negative triples ^−^ with the goal being to maximize the difference between their respective plausibility scores by a good margin λ. The hinge loss is computed using equation 5.

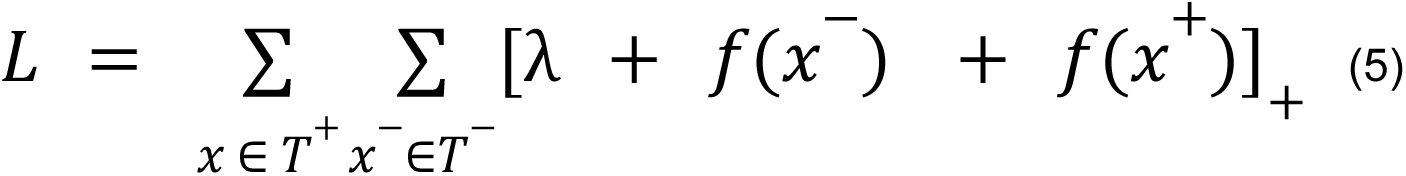

The node embeddings from a KGE model can be used for downstream applications such as node classification, link prediction etc.

#### 5.2.2 Downstreaming strategy to generate gene scores

In order to rank the genes in the geneset; the goal is to find the likelihood of the existence of a link between the gene nodes and the Alzheimer’s Disease nodes in the knowledge graph using the embeddings generated to represent the nodes and edges in the KG. The likelihood is quantified using a score that is generated by a scoring function that is specific to the KGE model [79]. The scoring function consumes embeddings for the head node (Alzheimer’s Disease node) and edge (relation) as input and assigns plausibility scores to several candidate tail nodes (gene nodes) based on the spatial proximity of the candidate gene node (tail) embeddings to that of the Alzheimer’s Disease node (head) embedding and the edge embedding that connects them. These plausibility scores are further normalized to bring them within the interval (0,1) and are used as gene scores from the knowledge graph. Scoring functions for various KGE models have been listed in Table 1.

For our use case; we leverage RotatE [83] to generate the node embeddings for Hetionet over other KGE models. The rationale for doing so has been explained in the results and discussion section of this paper. The node embeddings generated by RotatE on Hetionet are then used downstream to rank the gene nodes and assign a gene score for every gene in the geneset using the scoring function of RotatE by link prediction.

### 5.3 Our co-optimized learner

Our goal is to supplement the learning outcome of the deep graph infomax algorithm trained on experimental genetics data and PPI network with the learning outcome of a knowledge graph embedding model trained on hetionet and vice versa using the architecture described in Figure 5.

**Figure 5:**
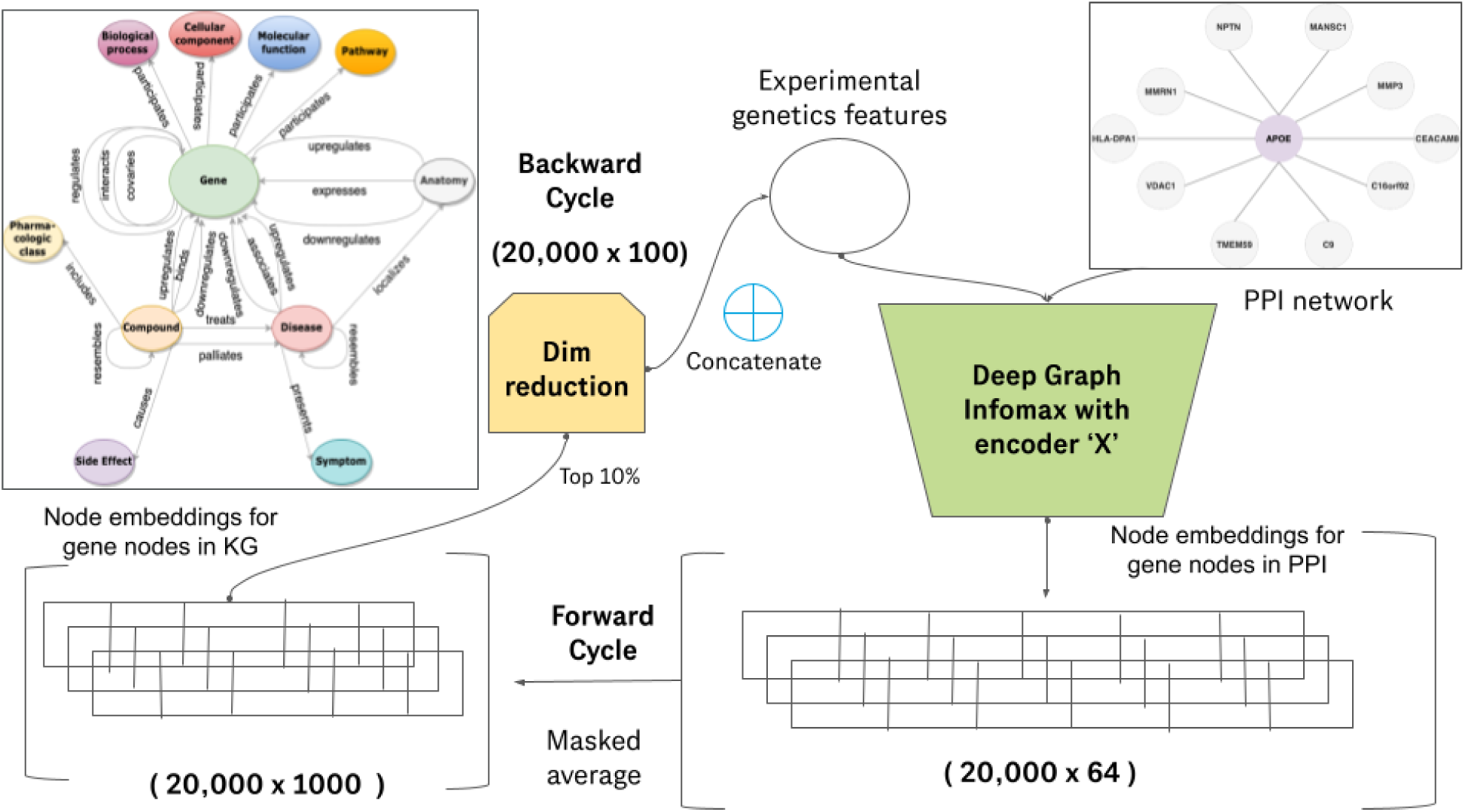
Architecture diagram of our co-optimized learner. One super epoch of training for our co-optimized learner consists of one forward cycle and one backward cycle. The learning outcome achieved by the deep graph infomax algorithm on the experimental genetics data and protein-protein interaction network is transferred to the knowledge graph embedding model during the forward cycle and the learning outcome achieved by the knowledge graph embedding model supplements the deep graph infomax algorithm in the backward cycle. Here, encoder X can be a Graph convolutional network (GCN), Graph attention network (GAT), GraphSAGE network, Predict and Propagate network (APPNP) or any other such graph representation learning algorithm.

### 5.4 Defining a super epoch

In order to achieve the same we need to propagate gene node embeddings from the deep graph infomax algorithm and incorporate it in the KGE model (forward cycle) and then we need to propagate the gene node embeddings from the KGE and incorporate it in the deep graph infomax algorithm (backward cycle). The forward cycle and backward cycle together constitute a super-epoch.

#### 5.4.1 Forward cycle

The gene node embeddings generated by the deep graph infomax algorithm are averaged with the gene node embeddings generated by the KGE model (RotatE) trained on hetionet. The dimensionality of the node embeddings from the deep graph infomax algorithm is much lesser than the dimensionality of the node embeddings from the KGE. Therefore we take a masked average of the two node embeddings and the dimension of the resultant node embedding vector is same as the dimension of the node embeddings of the RotatE model. Modifying the gene node embeddings of the KGE by augmenting it with the node embeddings of the deep graph infomax algorithm modifies the plausibility scores yielded by the scoring function of RotatE while generating link predictions. Therefore we get different sets of gene scores for the KGE model upon the completion of every forward cycle of a super epoch. The link predictions / ranked geneset produced as a consequence of the same are influenced by the experimental genetics data used to generate the deep graph infomax gene node embeddings.

#### 5.4.2 Backward cycle

The gene node embeddings generated by RotatE need to be incorporated into the deep graph infomax algorithm. The dimensionality of the node embeddings generated by the RotatE model on hetionet was pretty large (1000 dimensions). Therefore, we reduce the dimensionality by performing principal component analysis (PCA) [84] and select only the top 10% of the most informative dimensions in the node embedding vector. Thereafter, we concatenate these selected embedding dimensions alongside the experimental genetics data as node features in the protein-protein interaction network and train the next super-epoch of our algorithm. The node embeddings generated by the deep graph infomax algorithm are now not only influenced by the experimental genetics data but also by the biomedical literature data contained in the knowledge graph (Hetionet) as a result of this co-optimization strategy.

#### 5.4.3 Stopping criterion

A super epoch of co-optimization consists of one forward cycle and one backward cycle. We utilize an early stopping criterion and let our co-optimization algorithm run for a few number of super epochs until the unsupervised loss function given in equation 1 does not get minimized any further during the forward cycle of the co-optimization phase. This indicates that the learning outcome from the knowledge graph embedding model (Implication of causal genes based on evidence in literature) used to supplement the deep graph infomax algorithm has reached saturation, leaving no room for any further co-optimization.

### 5.5 Mutually informative score

After we complete the training of our algorithm; we have two sets of node embeddings for every gene in the geneset. One of those two sets of node embeddings is generated by the deep graph infomax algorithm and the other set of node embeddings is generated by the knowledge graph embedding model. We use separate downstreaming strategies for the node embeddings from the deep graph infomax algorithm and the node embeddings from the knowledge graph embedding algorithm as described in sections 5.1.2 and 5.2.2 in order to generate two components of gene score for every gene in the geneset. After generating the two components of gene score for every gene, we take an average of those two components for each gene to come up with the mutually informative score for each gene based on which we sort all the genes in the geneset to rank them.

### 5.6 Interpretability of the top ranked genes using saliency maps

Apart from generating the ranked geneset for target prioritization, we also want to understand which features from the experimental genetics data have been implicated by the algorithm to come up with the current set of rankings. We do this by leveraging the concept of integrated gradients in a geometric learning setting. Let us consider a node *G* in a graph. The node _1_ embeddings for node *G* is influenced by the node features of the nodes that can be reached 1 from *G* within 2 edges. The subnetwork that contains the nodes whose features influence the 1 node embeddings of node *G* is called the ego-net of *G*. Firstly; we start with a baseline / zero 1_1_ graph and then progressively add node *G* and other gene nodes connected to *G* along with 1_1_ their respective features and the links/edges that connects them together to the baseline graph in steps of alpha to create a sequence of graphs leading up to the full ego-net of the node *G*. _1_ Additionally, we also mask each node feature of *G* exactly once to observe the influence of a _1_ specific node feature on the predictions / gene scores. We compute the gradient for the graphs generated at each step as shown in Figure 6 to measure the relationship between changes to a feature and the corresponding changes in the model’s predictions / gene scores. Thereafter, a numerical approximation is computed by averaging the gradients.

**Figure 6:**
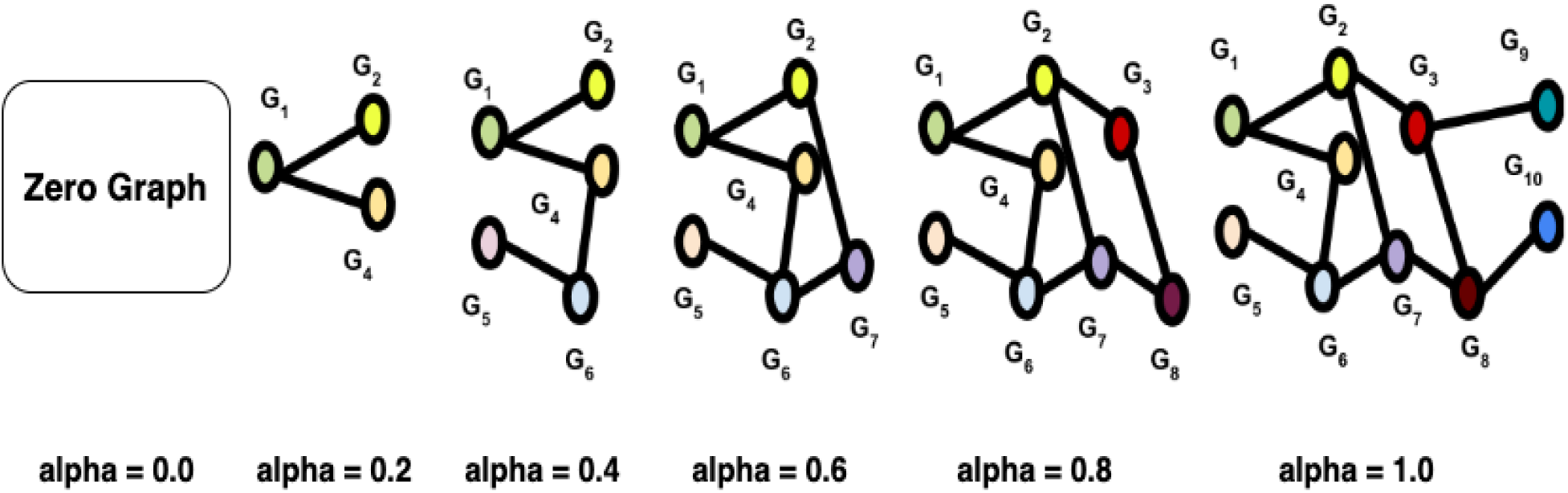
Interpretability for the gene score/ranking of a given gene G_1_ is achieved using saliency maps and integrated gradients. The nodes and edges neighboring G_1_ are progressively added to G_1_. The node feature importance, integrated node importance and link importance metrics are computed at every step based on the gradient change observed at that step upon adding a few nodes and edges from G_1_’s ego-net. The nodes and edges added in the steps corresponding to high gradient change are designated more important over other nodes and edges in the G_1_’s ego-net.

The result of this yields three important metrics to explain the gene scores and thereby the ranking of genes in terms of the features from the experimental genetics dataset and linkage in the PPI network. The three important metrics are:

a. Node feature importance: Change in the predicted gene score for a gene node when a particular node feature F of node *G* is masked.
b. Integrated node importance: The summation of feature importances for node *G* for all _1_ its node features.
c. Link importance: The change in gene score of *G* if a particular link is removed from 1 node *G*_1_ ego-net. 1

Once we identify the node feature importance, integrated node importance and link importance for the gene *G*_1_; we can explain the score and ranking of *G*_1_ assigned by our co-optimized model 1 in terms of those three metrics. Such a method of axiomatic attribution is known as *Integrated Gradients* [85][86].

Thereafter, we visualize the ego-net of *G*_1_ using a saliency map [87] with appropriate color coding for the gene nodes and edges in *G*_1_’*s* ego-net based on their importance in assigning the 1 score/ranking for *G*_1_ by the co-optimized learner.

### 5.7 Evaluating the performance of our algorithm

We evaluate the gene rankings generated from the embeddings of the individual algorithms that take part in co-optimization separately as well as collectively. We use rank based evaluation metrics to evaluate the gene rankings assigned by both the KGE as well as the deep graph infomax model.

#### 5.7.1 Calculation of rank based evaluation metrics for the gene rankings generated by co-optimized embeddings from KGE model

We evaluate the gene rankings generated by the co-optimized knowledge graph embeddings by computing two rank-based evaluation metrics namely Mean rank (MR) and Hits@k [43]. For a given triple (h,r,t) where h represents the Alzheimer’s Disease node, t represents the genes in the full geneset and r represents the relations connecting them together; we can generate mean rank and Hits@k for the silver standard genes to observe the rankings of the silver standard genes in the context of the full geneset. Since silver standard genes have been collected from expert curated data sources and causal evidence that has implicated their relevance to Alzheimer’s Disease, they should be ideally ranked higher than the other genes in the full geneset. The rank-based evaluation metrics (Mean Rank and Hits@K) generated for the genes in the silver standard geneset helps us to mathematically validate the same. The mean rank (MR) and Hits@K metrics can be computed using equations 6 and 7 for the rankings yielded by a knowledge graph embedding model to the genes in the silver standard geneset.

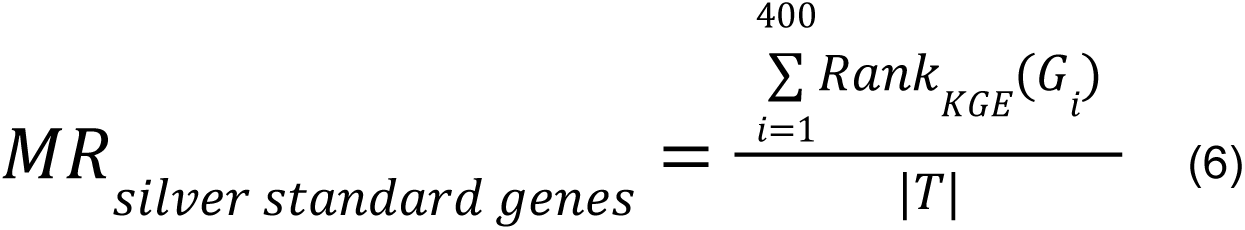

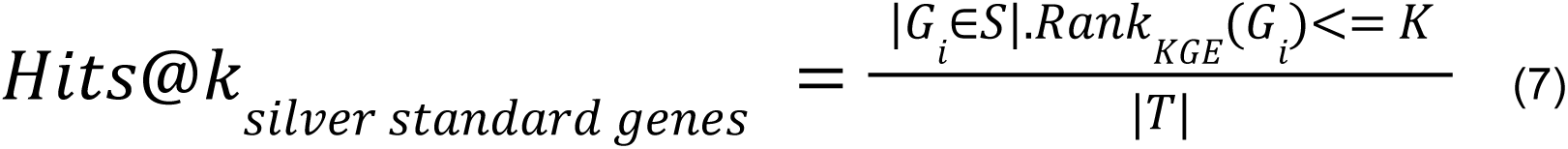

#### 5.7.2 Calculation of rank based evaluation metrics for the gene rankings generated by co-optimized embeddings from deep graph infomax model

The strategy that we have adopted for evaluating the gene rankings generated by the co-optimized embeddings from the deep graph infomax model is observing the mean rank of the silver standard genes (ranked purely using the gene scores from deep graph infomax algorithm) in the context of the fully ranked geneset. We computed the mean rank for the silver standard genes in the full geneset using equation 8.

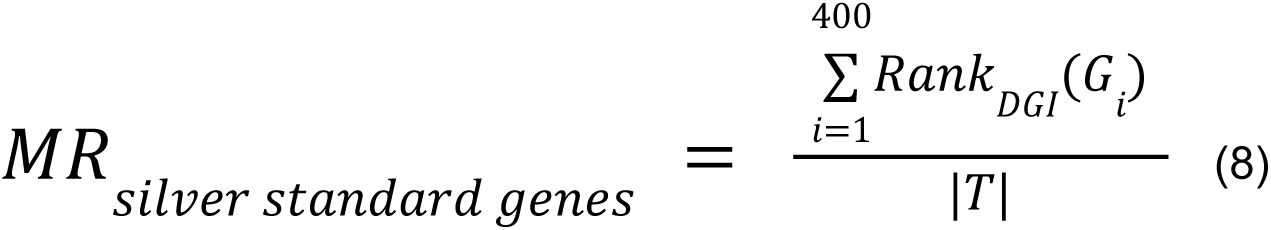

The silver standard genes should be ideally ranked higher than the remaining genes in the geneset since they have causal evidence associating them with Alzheimer’s Disease. The mean rank generated by doing this shows the capability of the deep graph infomax algorithm trained on experimental genetics data and protein-protein interaction network to rank the silver standard genes present in the full geneset.

#### 5.7.3 Calculating rank based evaluation metrics for the gene ranking generated by averaging gene scores from KGE and deep graph infomax algorithm

Similar to our strategy for evaluating the gene rankings / scores from individual models, we compute the mean rank for the silver standard genes in the fully ranked geneset that is sorted by the mutually informative scoring metric i.e. average of the gene scores obtained from the KGE model and the gene scores obtained from the deep graph infomax model for all the genes in the geneset using equations 9 and 10. The mean rank for the silver standard genes in the fully ranked geneset sorted by the mutually informative scoring metric helps us understand if supplementing the gene rankings from one domain with the gene rankings from another domain improves the mean rank of the silver standard genes in the fully ranked geneset.

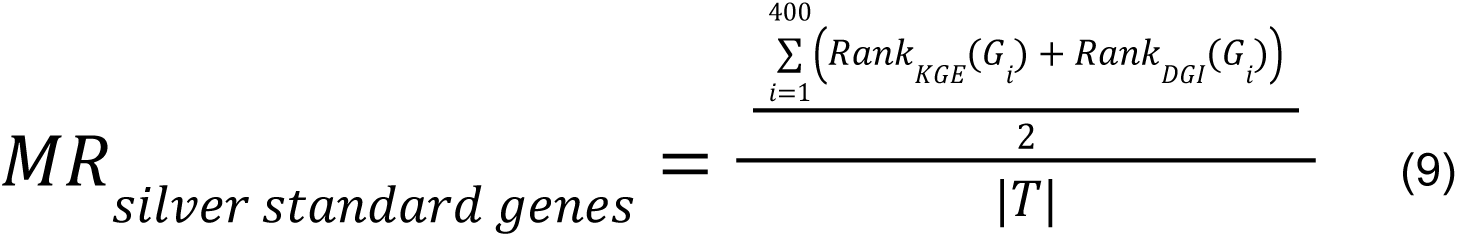

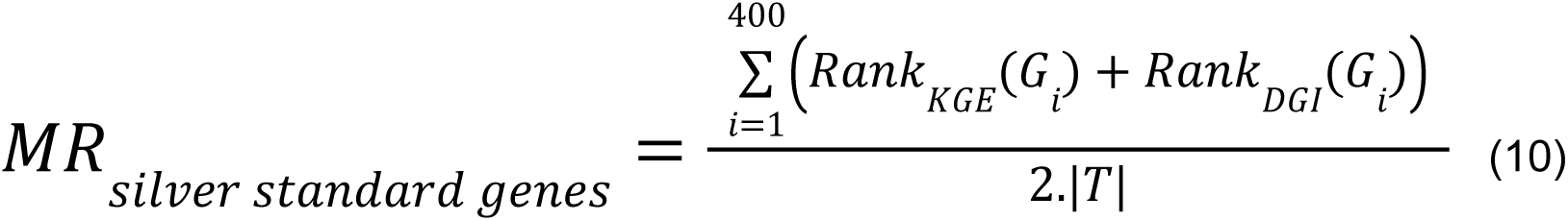

## 6. Experiment Setup

The knowledge graph embedding models in the following experiments were trained on hetionet using Pykeen [88] and the Deep graph infomax models were trained on the protein-protein interaction network using Stellargraph [89] on a high performance computing cluster that used a V100 Volta GPU with 32 GB of memory.

We conducted two different types of experiments where each experiment had three sub experiments within it as described below:

1. We evaluated the gene scores for the silver standard genes in the geneset by averaging the gene score assigned to a given silver standard gene by a KGE model (RotatE) and the gene score assigned to the same silver standard gene by a deep graph infomax model (with various GNN encoders) and generated a mean rank for all the silver standard genes in the geneset that is sorted and ranked by this average gene score. This experiment merely combines the individual scores from two separate models and does not co-optimize the gene embeddings during training.

a. We ranked the full geneset using the gene scores generated by the non co-optimized knowledge graph embedding model and obtained the mean rank for the silver standard genes in the context of the fully ranked geneset.
b. We ranked the full geneset using the gene scores generated by the non co-optimized deep graph infomax model and obtained the mean rank for the silver standard genes in the context of the fully ranked geneset.
c. We ranked the full geneset using the rankings generated by averaging the gene scores from the non co-optimized knowledge graph embedding model and the non co-optimized deep graph infomax model and obtained the mean rank for the silver standard genes in the context of the fully ranked geneset.
2. We evaluated the gene scores for the silver standard genes in the geneset by averaging the gene score assigned to a given silver standard gene by a co-optimized KGE model (RotatE) and the gene score assigned to the same silver standard gene by a co-optimized deep graph infomax model (with various GNN encoders) and generated a mean rank for all the silver standard genes in the geneset that is sorted and ranked by this average gene score (mutually informative score). This experiment uses co-optimized embeddings from KGE and deep graph infomax models to generate the averaged gene score / mutually informative score.

a. We ranked the full geneset using the gene scores generated by the co-optimized knowledge graph embedding model and obtained the mean rank for the silver standard genes in the context of the fully ranked geneset.
b. We ranked the full geneset using the gene scores generated by the co-optimized deep graph infomax model and obtained the mean rank for the silver standard genes in the context of the fully ranked geneset.
c. We ranked the full geneset using the rankings generated by averaging the gene scores from the co-optimized knowledge graph embedding model and the co-optimized deep graph infomax model and obtained the mean rank for the silver standard genes in the context of the fully ranked geneset.

Hyperparameter optimization:

Knowledge graph embedding model hyperparameters

- Embedding dimensions= [600, 800, 1000, 1200, 1400]
- Number of negative triples per positive triple = [1, 10, 100]
- Margin for margin ranking loss = [1, 4, 7, 10]
- Regularization coefficients = [0.02, 0.06, 0.10]

Deep graph infomax model hyperparameters

- GNN encoder layer sizes = [64, 128, 256]
- Number of GNN encoder layers = [1, 2, 3]
- Number of attention heads = [8, 10, 12] (Only if the GNN encoder is a GAT)
- Learning rate = [0.001, 0.005, 0.01]

Co-optimization hyperparameters:

Number of embedding dimensions from the KGE to be concatenated alongside the node features in the PPI network for the backward cycle of a super-epoch = [10%, 20%, 30%]

Early Stopping criterion:

In order to prevent the overfitting of our co-optimized learner we use early stopping to stop the training if the unsupervised loss function from the deep graph infomax algorithm does not find an even lower minima than the current minima after every 20 super-epochs of training.

Patience = 20

Lowest value observed for the validation loss remains the same even after 10 super-epochs.

## 7. Results and Discussion

Once we generate the co-optimized embeddings for the gene nodes we evaluate the gene scores assigned by our algorithm using those embeddings on a set of experiments to validate the following:

1. Averaging the gene scores from the KGE model and the gene scores from the deep graph infomax model yields a new set of gene scores for every gene that helps us rank the full geneset better.
2. Utilization of co-optimized embeddings during the process of training our algorithm has performance benefits over using non co-optimized embeddings from both the models separately.

Moreover, we need to experimentally check if we are indeed able to successfully induce a grouping around our silver standard genes as the gene scores assigned by our deep graph infomax embeddings leverages the spatial proximity of the embeddings of the non silver standard genes with that of the silver standard genes in the full geneset.

### 7.1 Result of clustering the co-optimized embeddings from deep graph infomax algorithm

The gene scores are assigned to each gene by the deep graph infomax algorithm on the basis of the proximity of the non-silver standard genes to the silver standard genes. Therefore, the gene score that is assigned to each gene by the deep graph infomax algorithm is a measure of proximity of its experimental genetic feature profile to that of the experimental genetic feature profile of a silver-standard gene. This creates inherent groups/clusters in the genesets that can be visualized by projecting the co-optimized gene embeddings to lower order dimensions using T-SNE [90]. The T-SNE clusters for all the co-optimized algorithms have been shown in Figure 7. Even in as few as two dimensions we can observe a clear grouping among the feature profiles of the genes in the geneset. This indicates that the embeddings of the non-silver standard genes are closely oriented to the embeddings of one or more silver standard genes in the latent space as represented by our algorithm. The grouping seems more so prominent in the case of a RotatE model that is co-optimized with a deep graph infomax model that uses a GCN encoder and a RotatE model that is co-optimized with a deep graph infomax model that uses a APPNP encoder as shown in Figure 7.

**Figure 7:**
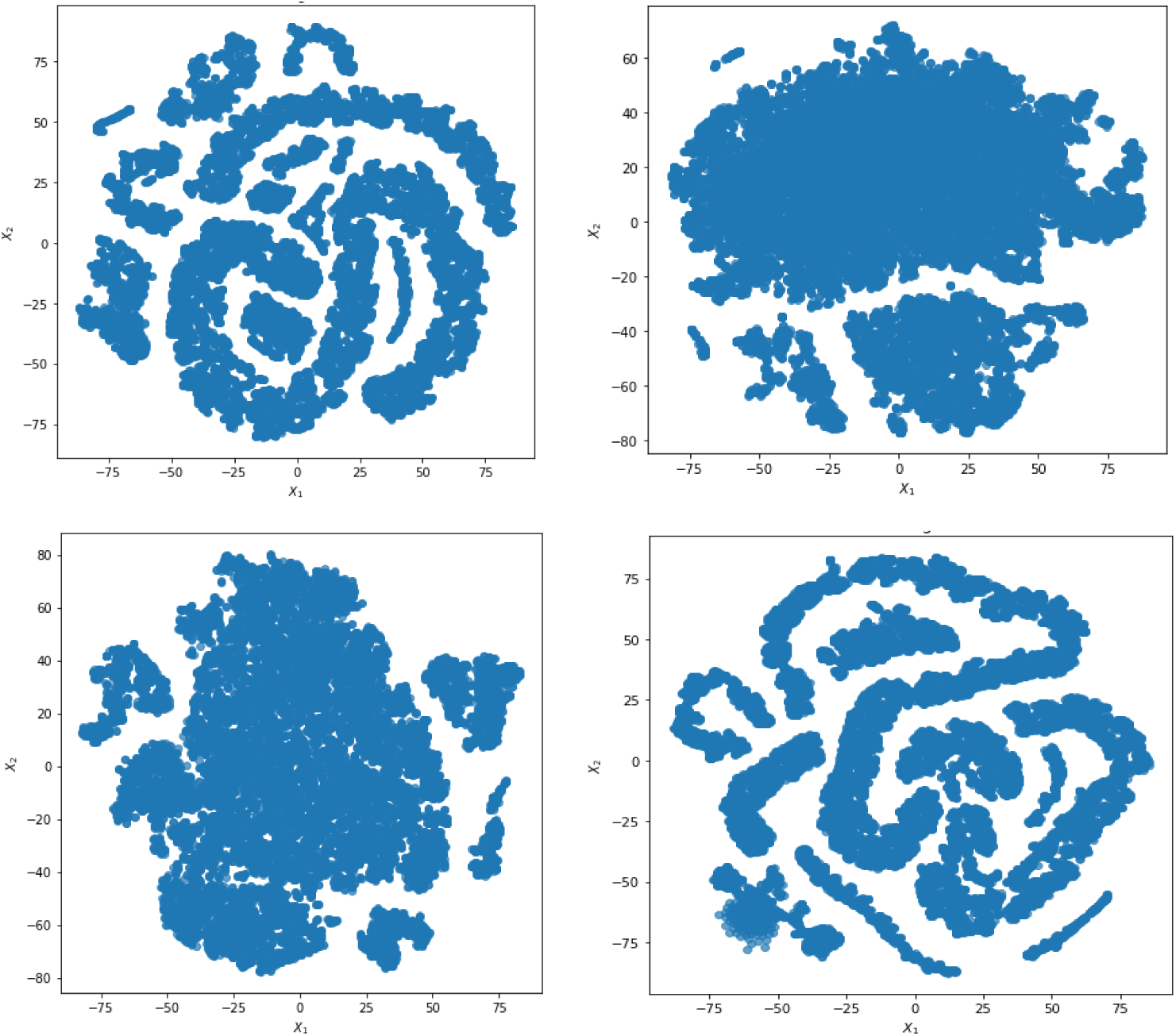
T-SNE clusters for visualizing the gene embeddings generated by four co-optimized gene scoring algorithms that use different GNN encoders for the deep graph infomax algorithm (a) RotatE + DGI (GCN) (b) RotatE + DGI (GAT) (c) RotatE + DGI(GraphSAGE) (d) RotatE + DGI (APPNP)

From the T-SNE grouping we can conclude that ranking all the genes based on their proximity to the silver standard genes helps us to induce an ordering based on the experimental similarity of a given gene with another gene that has known causal evidence implicating it with Alzheimer’s Disease.

### 7.2 Ablation studies

In order to observe the effect of co-optimization we conducted two different experiments:

1. We evaluated the gene scores for the silver standard genes in the geneset by averaging the gene score assigned to a given silver standard gene by a KGE model (RotatE) and the gene score assigned to the same silver standard gene by a deep graph infomax model (with various GNN encoders) and generated a mean rank for all the silver standard genes in the geneset that is sorted and ranked by this average gene score. This experiment merely combines the individual scores from two separate models and does not co-optimize the gene embeddings during training.
2. We evaluated the gene scores for the silver standard genes in the geneset by averaging the gene score assigned to a given silver standard gene by a co-optimized KGE model (RotatE) and the gene score assigned to the same silver standard gene by a co-optimized deep graph infomax model (with various GNN encoders) and generated a mean rank for all the silver standard genes in the geneset that is sorted and ranked by this average gene score (mutually informative score). This experiment uses co-optimized embeddings from KGE and deep graph infomax models to generate the averaged gene score / mutually informative score.

In order to verify the performance gain achieved by co-optimization; the mean rank of the silver standard genes in the geneset ranked by the co-optimized models should be lower than the mean rank of the silver standard genes in the geneset ranked by the non co-optimized models.

#### 7.2.1 Evaluating the gene scores averaged from KGE and deep graph infomax algorithm without co-optimization of the gene embeddings

Firstly, we generated mean rank for the silver standard genes using various standard knowledge graph embedding models [65][83][91][92] trained on hetionet and selected the knowledge graph embedding model with the lowest mean rank for the silver standard genes. Then we generated mean rank for the silver standard genes using the deep graph infomax algorithm trained purely on the protein-protein interaction network and the experimental genetics data that is incorporated as node features in the protein-protein interaction network. We combined the separate sets of gene scores from both these models by taking an average of the gene scores corresponding to each gene and then computed the mean rank for the silver standard genes in the geneset that is ranked/sorted by these average gene scores. It is important to note that the two models here score the genes independently and their respective gene embeddings are not co-optimized. Figure 8 showcases the mean rank of silver standard genes ranked by the KGE algorithm as well as the mean rank of silver standard genes ranked by the deep graph infomax algorithm in the absence of co-optimization.

**Figure 8:**
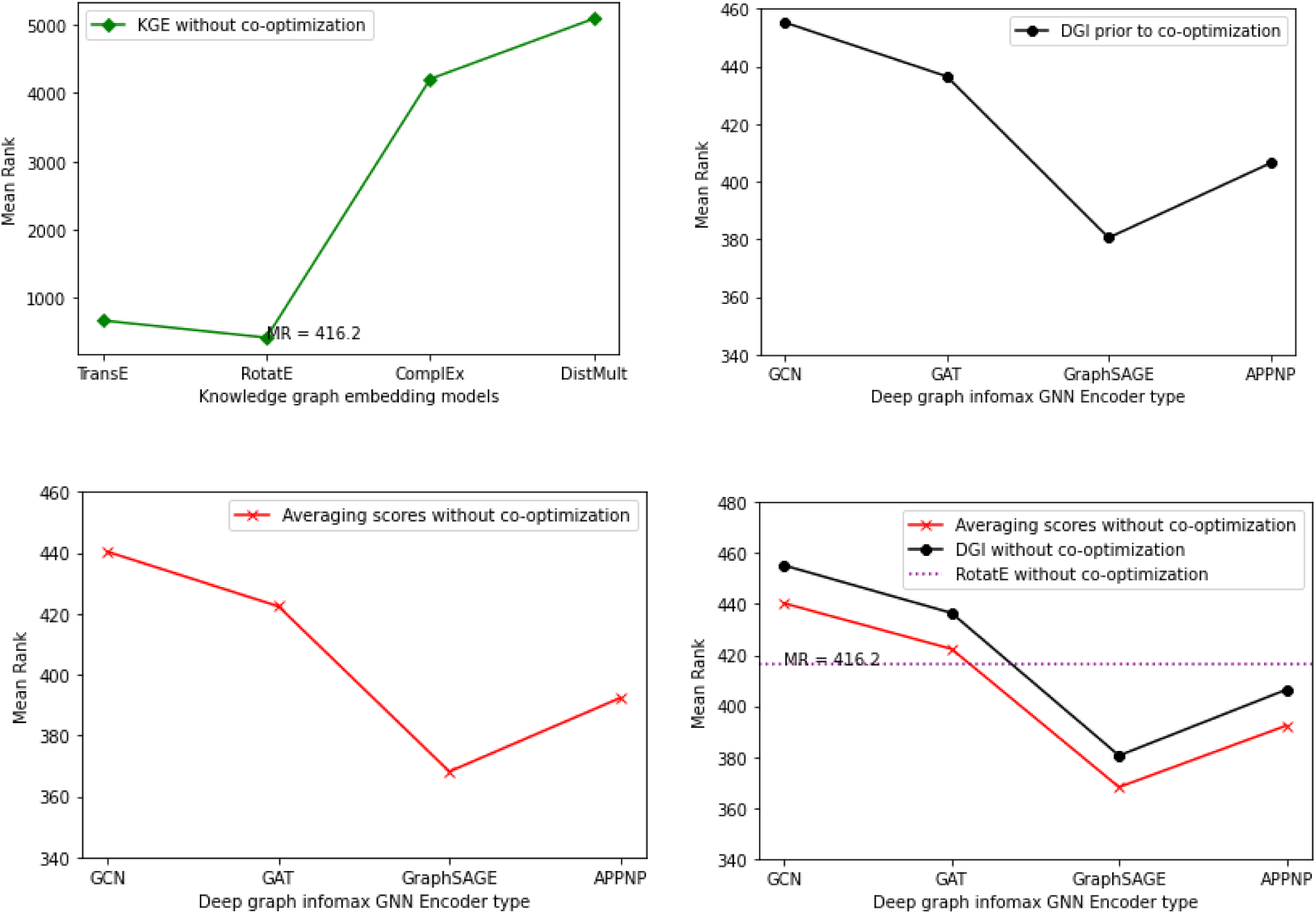
Plots showing the mean rank of the silver standard genes in the fully ranked geneset by non co-optimized KGE and deep graph infomax algorithms (a) Mean rank for silver standard genes from different KGE models (b) Mean rank for silver standard genes from non co-optimized deep graph infomax algorithm using different GNN encoders (c) Mean rank for silver standard genes whose gene scores are generated by averaging gene scores from KGE and deep graph infomax algorithm. (d) Mean rank for silver standard genes from RotatE, mean rank for silver standard genes from non co-optimized deep graph infomax algorithm and mean rank for silver standard genes whose gene scores are generated by averaging gene scores from KGE and deep graph infomax algorithm.

From Table 2, we observe that RotatE has the lowest mean rank and highest Hits@k for the silver standard genes in the fully ranked geneset among other standard KGE models. Therefore, we consider the gene scores derived from RotatE over the other KGE models. Likewise, using a GraphSAGE encoder for the deep graph infomax algorithm yields the lowest mean rank for the silver standard genes in the fully ranked geneset as shown in Table 3.

**Table 2.**
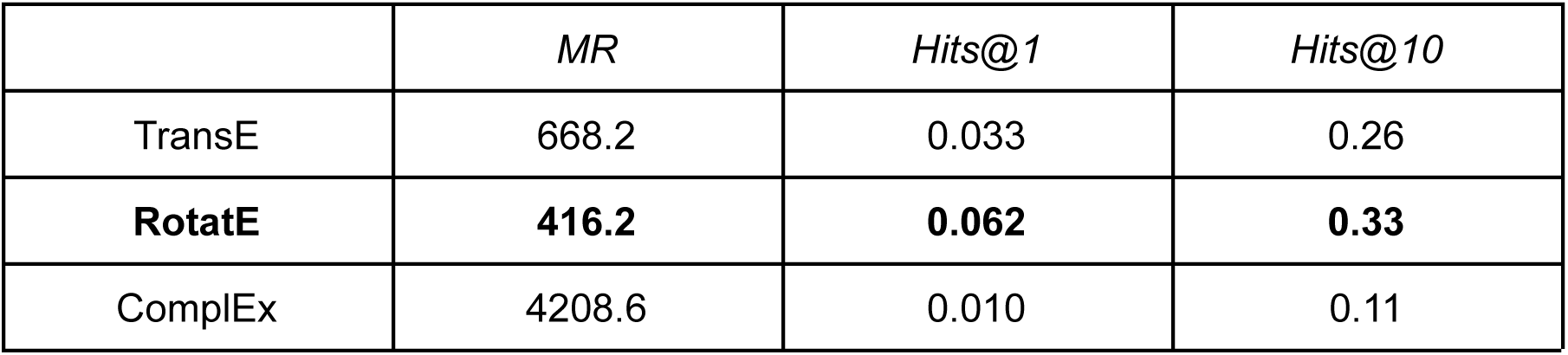
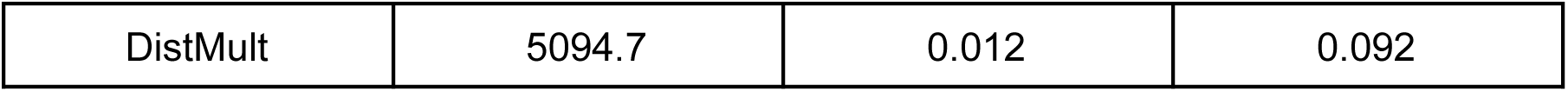
Rank-based evaluation metrics were computed for silver standard genes in the geneset that were ranked using standard knowledge graph embedding models.

**Table 3.** Mean rank for the silver standard genes were computed using different encoders in the deep graph infomax algorithm

| *Non co-optimized Deep Graph Infomax* | *MR* |
| --- | --- |
| GCN | 455.24 |
| GAT | 436.62 |
| **GraphSAGE** | **380.52** |
| APPNP | 406.44 |

Ranking the silver standard genes using the average of the gene scores from RotatE and deep graph infomax algorithm boosts the mean rank of the silver standard genes in the context of the fully ranked geneset as shown in Table 4. The aggregated gene score generated by averaging the gene scores from RotatE and GraphSAGE yielded the lowest mean rank for the silver standard genes amongst the other models shown in Table 4.

**Table 4.** Mean rank for silver standard genes whose gene scores were generated by averaging the gene scores from KGE and deep graph infomax algorithms.

| *Averaging gene scores without co-optimization* | *MR* |
| --- | --- |
| RotatE | 416.2 |
| RotatE + GCN | 440.34 |
| RotatE + GAT | 422.4 |
| **RotatE + GraphSAGE** | **368.12** |
| RotatE + APPNP | 392.32 |

#### 7.2.2 Evaluating the gene scores averaged from KGE and deep graph infomax algorithm with co-optimization of the gene embeddings

We ranked the full geneset using the rankings generated by averaging the gene scores from the co-optimized knowledge graph embedding model and the co-optimized deep graph infomax model and obtained the mean rank for the silver standard genes in the context of the fully ranked geneset. The purpose of conducting this experiment was to observe the impact on rank based evaluation metrics in the presence and absence of co-optimization. Figure 9 showcases the effect on the mean rank of silver standard genes for both KGE as well as the deep graph infomax algorithm prior to co-optimization and post co-optimization.

**Figure 9:**
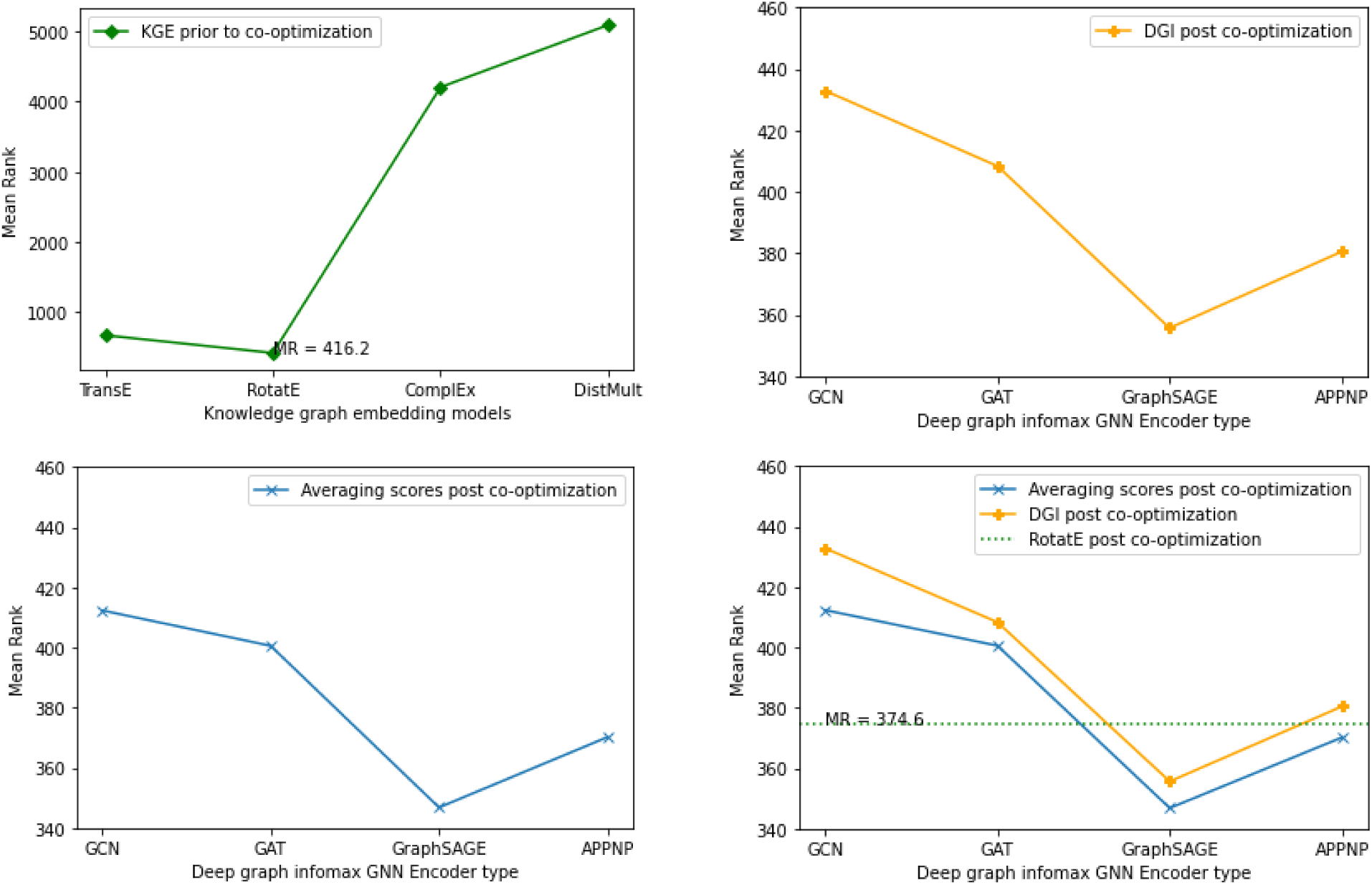
Plots showing the mean rank of the silver standard genes in the fully ranked geneset by co-optimized KGE and deep graph infomax algorithms (a) Mean rank for silver standard genes from different KGE models prior to co-optimization (b) Mean rank for silver standard genes from the deep graph infomax algorithm using different GNN encoders prior to co-optimization (c) Mean rank for silver standard genes whose gene scores are generated by the deep graph infomax algorithm post co-optimization with RotatE. (d) Mean rank for silver standard genes from co-optimized RotatE, mean rank for silver standard genes from deep graph infomax algorithm prior to co-optimization and mean rank for silver standard genes whose gene scores are generated by the deep graph infomax algorithm post co-optimization with RotatE

We generated mean rank for the silver standard genes using RotatE after supplementing it with node embeddings from the deep graph infomax algorithm through co-optimization. The rank based evaluation metrics (Mean rank and Hits@k) showed significant improvement for RotatE post co-optimization when compared to the rank based evaluation metrics observed in the absence of co-optimization as shown in Table 2 and Table 5. Likewise, the mean rank of silver standard genes in the geneset ranked by the co-optimized deep graph infomax algorithm shown in Table 6 reduced in comparison to the mean rank of silver standard genes in the geneset ranked by the deep graph infomax algorithm in the absence of co-optimization as shown in Table 3. This indicated that both the KGE algorithm and the deep graph infomax algorithm benefitted from the co-optimization procedure as validated by the improvement observed in the mean rank of the silver standard genes in the fully ranked geneset.

**Table 5.** Rank-based evaluation metrics were computed for silver standard genes in the geneset that was ranked using the co-optimized RotatE model.

|  |  |  |  |
| --- | --- | --- | --- |
|  | *MR* | *Hits@1* | *Hits@10* |
| **RotatE** | **374.6** | **0.092** | **0.36** |

**Table 6.** Mean rank for silver standard genes whose gene scores were generated by the deep graph infomax algorithm post co-optimization with RotatE.

| *Co-optimized Deep Graph Infomax* | *MR* |
| --- | --- |
| GCN | 432.82 |
| GAT | 408.36 |
| **GraphSAGE** | **355.64** |
| APPNP | 380.46 |

Similar to the boost in mean rank observed when ranking the silver standard genes using the average of the gene scores from non co-optimized RotatE and deep graph infomax algorithms in Table 4, ranking the silver standard genes using the average of the gene scores from co-optimized RotatE and deep graph infomax algorithms also boosts the mean rank of the silver standard genes in the context of the fully ranked geneset as shown in Table 7. From Table 4 and Table 7 it is evident that averaging the gene scores from the deep graph infomax model and the knowledge graph embedding model further improves the mean rank of the silver standard genes in the fully ranked geneset.

**Table 7.** Mean rank for silver standard genes whose gene scores were generated by averaging the gene scores from co-optimized KGE and deep graph infomax algorithms.

| *Averaging gene scores post co-optimization* | *MR* |
| --- | --- |
| RotatE | 374.60 |
| RotatE + GCN | 412.35 |
| RotatE + GAT | 400.58 |
| **RotatE + GraphSAGE** | **346.81** |
| RotatE + APPNP | 370.21 |

#### 7.2.3 Comparison between combining gene scores from RotatE and Deep graph infomax models in the presence and absence of co-optimization

By juxtaposing the mean rank plots in the presence and absence of co-optimization as shown in Figure 10, it is evident that the effect of co-optimizing the node embeddings from the deep graph infomax algorithm and the RotatE algorithm (knowledge graph embedding algorithm) improves the mean rank of the silver standard genes and furthermore averaging the gene scores from the co-optimized models and ranking the geneset based on the average score yields the best mean rank for the silver standard genes in the geneset.

**Figure 10:**
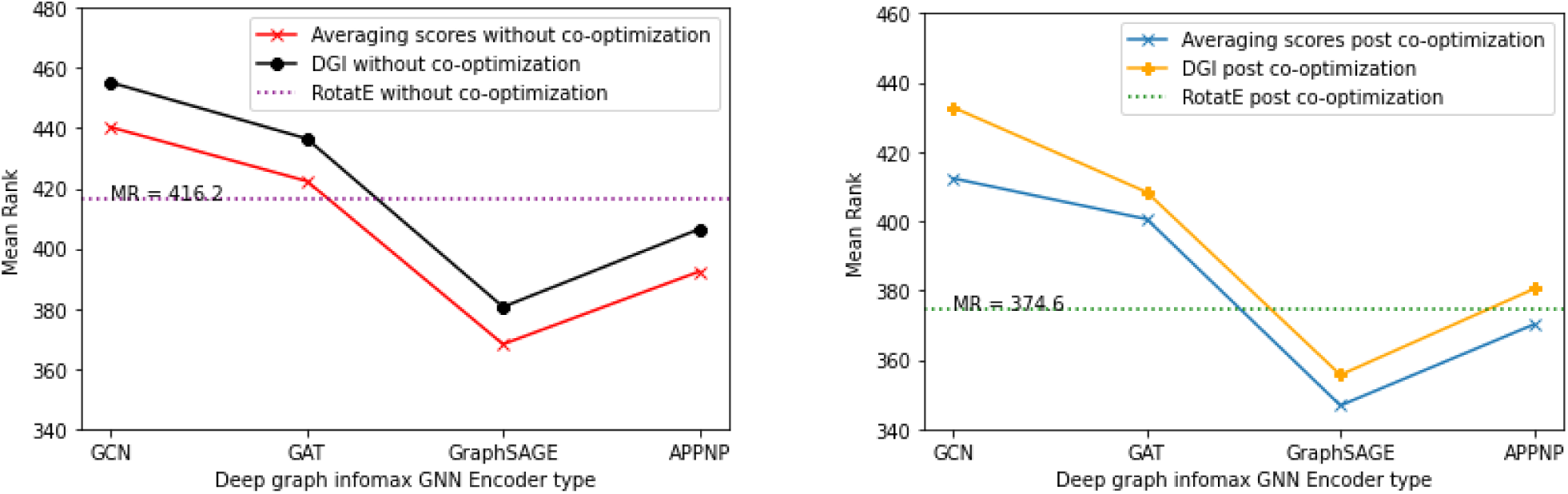
Comparison of the mean ranks for the silver standard genes with and without co-optimization. (a) Mean rank for the silver standard genes ranked by the non co-optimized KGE and deep graph infomax algorithm. (b) Mean rank for the silver standard genes ranked by the co-optimized KGE and deep graph infomax algorithm.

### 7.3 Explaining the gene rankings generated by our algorithm

In order to examine the most relevant features of the experimental data that are responsible for the implication of a given gene as being causal to AD and thereby getting ranked at the top of the prioritized geneset by our co-optimized model; some of the top ranked silver standard genes by our best performing co-optimized algorithm (RotatE + Deep Graph Infomax with a GraphSAGE encoder) have been considered below:

1. PDGFRB (Platelet derived growth factor receptor beta)
2. CYBB (Cytochrome b-245 beta chain)
3. KIF5A (Kinesin Family Member 5A)
4. VDR (Vitamin D Receptor)
5. ARAF (A-Raf proto-oncogene)
6. APOE (Apolipoprotein E)

As shown in Figure 11, we utilize saliency maps to visualize the node feature importance, integrated node importance and link importance in a given gene’s ego-net. The interpretability metrics such as node feature importance, integrated node importance andlink importance are computed using integrated gradients by fitting a GNN model that uses the gene scores as the labels for the respective nodes in the protein-protein interaction network. Let us consider the top 6 ranked silver standard genes to examine the node feature importance given to the experimental genetic node features corresponding to the genes, node importance given to other connected genes and the link importance given to the edge connecting the genes together in the protein-protein interaction network.

**Figure 11:**
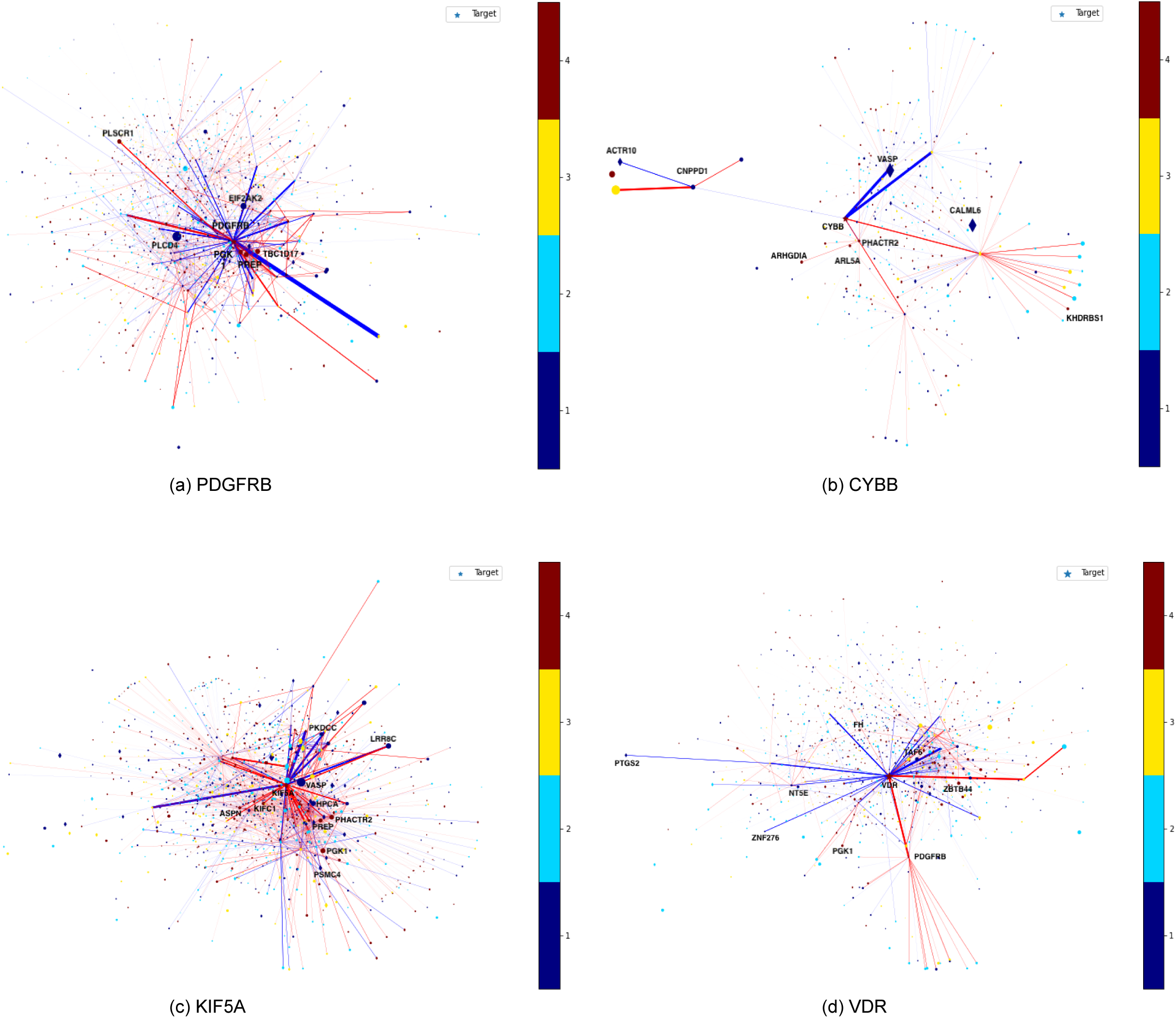
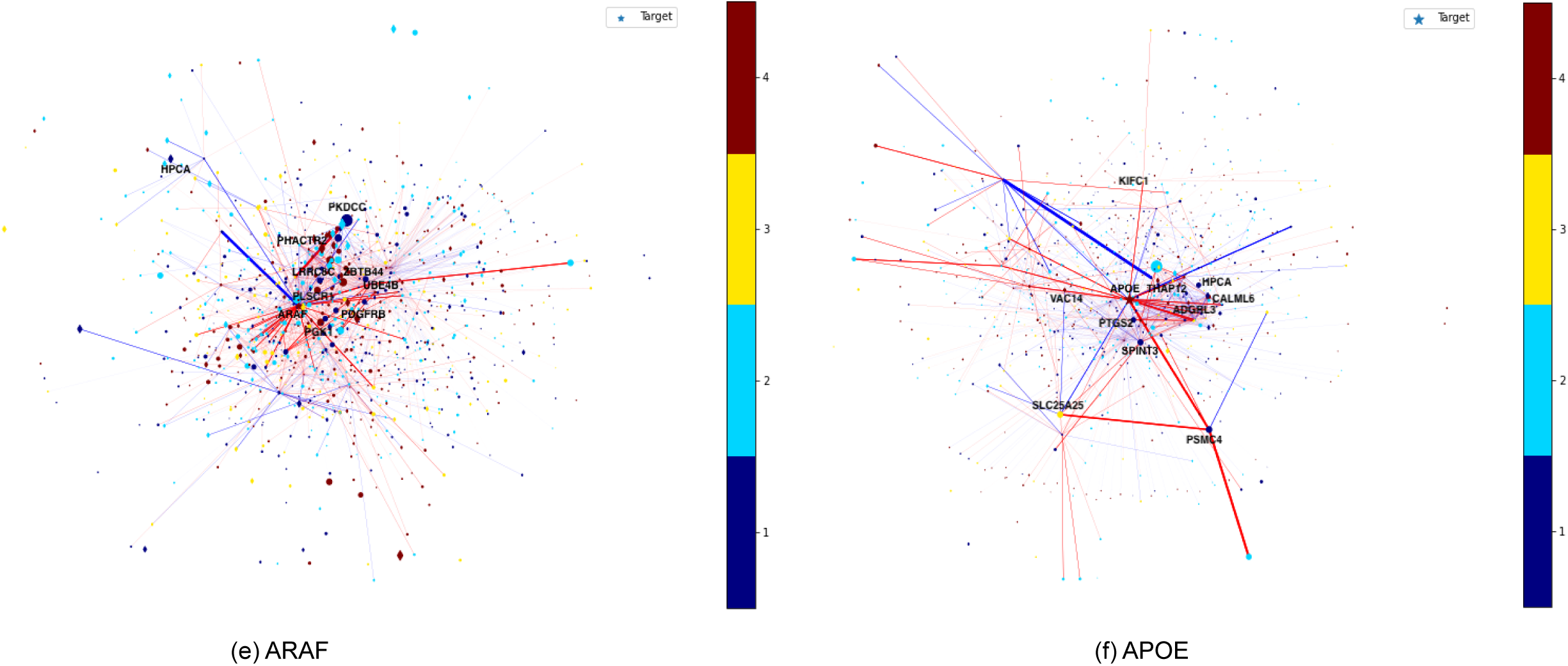
Saliency maps that showcase the important genes and protein-protein interactions that impact the gene score assigned to each of the top ranked silver standard gene by our algorithm (a) PDGFRB (b) CYBB (c) KIF5A (d) VDR (e) ARAF and (f) APOE. The red colored nodes and edges indicate higher importance in determining the gene score of the target silver standard gene and the blue colored nodes and edges indicate lower importance in determining the gene score of the target silver standard gene. A bigger sized node in the saliency map has more impact on the gene score of the target silver standard gene than a smaller sized node. Diamond shaped nodes have greater impact on the gene score of target silver standard gene than circular nodes.

PDGFRB (Platelet derived growth factor receptor beta) Node feature importance:

1. Risk of developing Alzheimer’s Disease based on its prevalence in family history (Common variant analysis feature)
2. Differential gene expression of PDGFRB for multiple cell types (microglia, astrocyte and neuron) in a transgenic mouse model of Alzheimer’s Disease (AD) where the mice were sacrificed after 13 months when amyloid pathology and microgliosis became pervasive. (Expression analysis feature)
3. Differential gene expression of PDGFRB for choroid plexus cells in a triple transgenic mouse model of Alzheimer’s Disease (AD) measured using single cell RNA sequencing from tissue samples of the hippocampus. It identifies the transcriptional response associated with amyloid and tau pathology in choroid plexus cells. (Expression analysis feature)
4. Differential gene expression of PDGFRB in the brain tissue microglia from the prefrontal cortex collected using single nucleus RNA sequencing. (Expression analysis feature)
5. Presence of rare variants in PDGFRB that can alter the age of onset of Alzheimer’s Disease (AD) detected using Whole Exome Sequencing (WES) and gene burden test. (Rare variant analysis feature)

Integrated node importance: PDGFRB, TBC1D17, PGK1, PREP, PLSCR1, THAP12, CLN8, SAAL1, DUSP13, TMEM9

Link importance:

- PDGFA, GABARAP
- PDGFRB, HIP1
- PDGFRB, PTPRS
- PDGFRB, SH3KBP1
- HIP1, ACTC1

Signaling Pathway activation:

- Apoptotic Pathways in Synovial Fibroblasts
- GPCR Pathway
- ERK Signaling
- PI3K/AKT Signaling

Molecular function: Developmental protein, Receptor, Transferase, Tyrosine-protein Kinase Biological process: Chemotaxis

CYBB (Cytochrome b-245 beta chain)

Node feature importance:

1. Differential gene expression of CYBB for endothelial cells in a triple transgenic mouse model of Alzheimer’s Disease (AD) measured using single cell RNA sequencing from tissue samples of the hippocampus. It identifies the transcriptional response associated with amyloid and tau pathology in endothelial cells. (Expression analysis feature)
2. Differential gene expression of CYBB for excitatory neurons in a triple transgenic mouse model of Alzheimer’s Disease (AD) measured using single cell RNA sequencing from tissue samples of the hippocampus. It identifies the transcriptional response associated with amyloid and tau pathology in excitatory neurons. (Expression analysis feature)
3. Differential gene expression of CYBB for astrocytes in Alzheimer’s Disease (AD) patients measured using sorted cell RNA sequencing from tissue samples of the superior frontal gyrus in the frontal cortex. (Expression analysis feature)
4. Differential gene expression of TREM2 gene in microglia response to amyloid-beta deposition in an Alzheimer’s Disease (AD) mouse model named 5XFAD. (Expression analysis feature)
5. Differential gene expression of CYBB in the oligodendrocyte precursor cells from the prefrontal cortex collected using single nucleus RNA sequencing. (Expression analysis feature)

Integrated node importance:

PHACTR2, ARL5A, CYBB, ARHGDIA, KHDRBS1, PAK5, RNF19A, DAAM2, NCKAP1, ACTR5

Link importance:

- CYBC1, P2RX1
- CYBB, GNG8
- CYBC1, CD79B
- CYBB, SOD1
- CYBC1, AQP10

Signaling Pathway activation:

- Signaling by Rho GTPases
- Innate Immune System
- Class I MHC mediated antigen processing
- Antigen processing-Cross presentation

Molecular function: Ion channel, Oxidoreductase, Voltage-gated channel

Biological process: Electron and Ion transport

<u>KIF5A</u> (Kinesin Family Member 5A)

Node feature importance:

1. Differential gene expression of KIF5A in oligodendrocytes from the prefrontal cortex collected using single nucleus RNA sequencing. (Expression analysis feature)
2. Differential gene expression of TREM2 gene in microglia response to amyloid-beta deposition in an Alzheimer’s Disease (AD) mouse model named 5XFAD. (Expression analysis feature)
3. Differential gene expression of KIF5A for astrocytes in Alzheimer’s Disease (AD) patients measured using sorted cell RNA sequencing from tissue samples of the superior frontal gyrus in the frontal cortex. (Expression analysis feature)
4. Differential gene expression of KIF5A for excitatory neurons in a triple transgenic mouse model of Alzheimer’s Disease (AD) measured using single cell RNA sequencing from tissue samples of the hippocampus. It identifies the transcriptional response associated with amyloid and tau pathology in excitatory neurons. (Expression analysis feature)
5. Differential gene expression of KIF5A for astrocytes in a triple transgenic mouse model of Alzheimer’s Disease (AD) measured using single cell RNA sequencing from tissue samples of the hippocampus. It identifies the transcriptional response associated with amyloid and tau pathology in astrocytes. (Expression analysis feature)

Integrated node importance:

PGK1, PHACTR2, PREP, ASPN, KIFC1, ARL5A, TMEM9, ERMP1, PDE3A, NME7

Link importance:

- PTDSS2, PTDSS2
- PFN4, ACTC1
- KIF5A, DYNLL2
- NPEPPS, UBA52
- KIF5A, TRAK2

Signaling Pathway activation:

- Golgi-to-ER retrograde transport
- Signaling by Rho GTPases
- Vesicle-mediated transport
- Class I MHC mediated antigen processing

Molecular function: Motor Protein

Biological process: Anterograde axonal protein transport, Chemical synaptic transmission and Microtubule-based movement

<u>VDR</u> (Vitamin D Receptor)

Node feature importance

1. Differential gene expression of VDR for choroid plexus cells in a triple transgenic mouse model of Alzheimer’s Disease (AD) measured using single cell RNA sequencing from tissue samples of the hippocampus. It identifies the transcriptional response associated with amyloid and tau pathology in choroid plexus cells. (Expression analysis feature)
2. Differential gene expression of VDR in oligodendrocytes from the prefrontal cortex collected using single nucleus RNA sequencing. (Expression analysis feature)
3. Differential gene expression of TREM2 gene in microglia response to amyloid-beta deposition in an Alzheimer’s Disease (AD) mouse model named 5XFAD. (Expression analysis feature)
4. Differential gene expression of VDR for Cajal-Retzius cells in a triple transgenic mouse model of Alzheimer’s Disease (AD) measured using single cell RNA sequencing from tissue samples of the hippocampus. It identifies the transcriptional response associated with amyloid and tau pathology in Cajal-Retzius cells. (Expression analysis feature)
5. Differential gene expression of VDR for astrocytes in Alzheimer’s Disease (AD) patients measured using sorted cell RNA sequencing from tissue samples of the superior frontal gyrus in the frontal cortex. (Expression analysis feature)

Integrated node importance:

PDGFRB, PGK1, ZBTB44, FH, PLSCR1, DPEP1, STX6, UBXN10, LMNA, AP1S2

Link importance:

- VDR, FA2H
- VDR, IL12B
- IL12B, IL23R
- IL12B, IL23A
- CCDC78, VDR

Signaling Pathway activation:

- Gene transcription
- Nuclear receptor transcription
- Metabolism of proteins
- Metabolism of steroids

Molecular function: DNA-binding, Receptor

Biological process: Transcription, Transcription regulation

<u>ARAF</u> (A-Raf proto-oncogene)

Node feature importance:

1. Differential gene expression of TREM2 gene in microglia response to amyloid-beta deposition in an Alzheimer’s Disease (AD) mouse model named 5XFAD. (Expression analysis feature)
2. Differential gene expression of ARAF for astrocytes in a triple transgenic mouse model of Alzheimer’s Disease (AD) measured using single cell RNA sequencing from tissue samples of the hippocampus. It identifies the transcriptional response associated with amyloid and tau pathology in astrocytes. (Expression analysis feature)
3. Risk of developing Alzheimer’s Disease based on evidence from UK Biobank GWAS catalog. (common variant analysis feature)
4. Differential gene expression of ARAF in microglial cells from the prefrontal cortex collected using single nucleus RNA sequencing. (Expression analysis feature)
5. Differential gene expression of ARAF for microglia in a transgenic mouse model of tauopathy. (Expression analysis feature)

Integrated node importance:

PDGFRB, PGK1, ZBTB44, PHACTR2, PLSCR1, DPEP1, ASPN, TMEM9, FAM153B, PDE3A

Link importance:

- MAP2K2, LSM1
- MAP2K1, LSM1
- GNA12, GNG5
- ARAF, LNX1
- GNL3, UBA52

Signaling Pathway activation:

- Prolactin signaling
- IL-9 signaling
- CNTF signaling
- GPCR signaling

Molecular function: Kinase, Serine/threonine-protein kinase, Transferase

Biological process: MAPK cascade, protein phosphorylation and modification, regulation of proteasomal ubiquitin dependent protein catabolic process, regulation of TOR signaling

APOE (Apolipoprotein E) Node feature importance:

1. Differential gene expression of APOE for microglia in a transgenic mouse model of tauopathy. (Expression analysis feature)
2. Differential gene expression of APOE for endothelial cells in a triple transgenic mouse model of Alzheimer’s Disease (AD) measured using single cell RNA sequencing from tissue samples of the hippocampus. It identifies the transcriptional response associated with amyloid and tau pathology in endothelial cells. (Expression analysis feature)
3. Differential gene expression of APOE in the oligodendrocyte precursor cells from the prefrontal cortex collected using single nucleus RNA sequencing. (Expression analysis feature)
4. Presence of rare variants in APOE that can alter the risk for Alzheimer’s Disease (AD) detected using Whole Exome Sequencing (WES) and gene burden test. (Rare variant analysis feature)
5. Differential gene expression of APOE for oligodendrocytes in a triple transgenic mouse model of Alzheimer’s Disease (AD) measured using single cell RNA sequencing from tissue samples of the hippocampus. It identifies the transcriptional response associated with amyloid and tau pathology in oligodendrocytes. (Expression analysis feature)

Integrated node importance:

VAC14, ADGRL3, THAP12, SLC25A25, KIFC1, ARL5A, TMCC2, HERPUD1, PLK4, NECTIN1

Link importance:

- APOC2, TTPA
- APOE, APOC2
- APOC2, APOA2
- APOA2, APOC2
- APOE, APOA1

Signaling Pathway activation:

- Plasma lipoprotein assembly, remodeling, and clearance
- Statin inhibition of cholesterol production
- Visual phototransduction
- Binding of ligands by scavenger receptors

Molecular function: Heparin binding

Biological process: Cholesterol metabolism, Host-virus interaction, Lipid metabolism, Lipid transport, Steroid metabolism, Sterol metabolism, Transport

## 8. Conclusion

Co-optimizing graph neural network embeddings with knowledge graph embeddings is a novel approach of incorporating prior knowledge from a large knowledge graph in the training procedure of a graph neural network algorithm on another domain of graph data with the goal being to supplement the learning outcome from the graph neural network with the prior knowledge from the knowledge graph and vice versa. For our use case to prioritize drug targets for Alzheimer’s Disease; supplementing the learning outcome from experimental data with that of the learning outcome from literature and viceversa proved to be an effective method in identifying causal genes relevant to Alzheimer’s Disease whose expression levels can be regulated to explore potential drug targets for treating Alzheimer’s Disease. The effectiveness of our strategy is validated by the low mean rank achieved for the silver standard genes in the context of the fully ranked geneset. Since the silver standard genes are known to be causal in nature; a low mean rank achieved for the silver standard genes when ranking all the genes in the geneset essentially entails that our algorithm is effective in prioritizing the causal genes for Alzheimer’s Disease based on the experimental data and literature evidence available for the same.

## Data Availability

The sources for all the data produced in the present work are cited in the manuscript.

http://www.nealelab.is/uk-biobank

https://www.ukbiobank.ac.uk/

https://www.ebi.ac.uk/gwas/

https://www.omim.org/

https://het.io/

https://go.drugbank.com/

https://www.ebi.ac.uk/chembl/

https://www.opentargets.org/

## Notes

### Competing Interest Statement

The authors have declared no competing interest.

### Funding Statement

Genentech Inc. and Roche Holding AG

### Author Declarations

The study used ONLY openly available human data from GWAS catalog, UK Biobank and the Hetionet knowledge graph.

